# Differentiating Mpox Infection and Vaccination Using a Validated Multiplex Orthopoxvirus IgG Serology Assay

**DOI:** 10.1101/2025.10.24.25338410

**Authors:** Jonathan C. Reed, Cecilia Downs, Kaden McAllister, Clarice Mauer, Christopher L. McClurkan, Donna Wilson, Kate Holzhauer, Jane A. Dickerson, Chase A. Cannon, Tara M. Babu, Matthew R. Golden, David M. Koelle, Alexander L. Greninger

## Abstract

The resurgence of monkeypox virus (MPXV) outbreaks has increased demand for validated serological assays to assess exposure and immunity. Cross-reactivity among orthopoxivuses, stemming from high sequence conservation, complicates distinguishing antibody responses from natural MPXV infection versus vaccination or other orthopoxvirus exposures. We validated the Meso Scale Discovery (MSD) V-PLEX Orthopoxvirus Panel 1 (IgG) Kit, which quantifies antibody levels to five MPXV antigens and their vaccina virus (VACV) orthologs, following Good Clinical Laboratory Practice (GCLP) guidelines. We assessed assay performance using serum from 26 individuals with prior mpox, 52 JYNNEOS vaccine recipients, and 179 unexposed controls. The assay reliably detected antibody responses in all exposed cohorts with peak levels observed 2-months post-vaccination. Antibody levels to specific antigens also correlated with Modified Vaccinia Ankara (MVA) neutralization titer, particularly for MPXV B6R/VACV B5R, MPXV E8L/VACV D8L, and MPXV M1R/VACV L1. Receiver operating characteristic (ROC) analysis showed that some individual antigens achieved high sensitivity and specificity for exposure detection (AUC > 0.96 for VACV D8L, MPXV B6R, VACV B5R); however, individual antigens performed poorly in distinguishing infection from vaccination. In contrast, antibody level ratios between some MPXV and VACV orthologs effectively differentiated MPXV infection from vaccinia vaccination with high sensitivity and specificity (e.g. MPXV A35R/VACV A33R ortholog ratio, AUC = 0.97, sensitivity = 0.97, specificity = 0.96). Our findings validate the MSD assay for clinical research and serosurveillance to assess MPXV immunity and support the utility of ortholog pair ratio analysis as a strategy to discriminate vaccinated and infected individuals.

**Importance:** Mpox continues to spread around the world, with recent data showing increasing incidence in the United States. While there are multiple FDA-authorized real-time PCR tests for diagnostic use, there are no FDA-authorized serological tests and few laboratory-developed serological tests offered. We evaluated the Meso Scale Discovery V-PLEX Orthopoxvirus Panel 1 (IgG) Kit according to Good Clinical Laboratory Practice guidelines and found that assay reliably detected antibody responses in mpox and vaccinia exposed cohorts and could distinguish them from unexposed cohorts. Intriguingly, we found that antibody level ratios between certain MPXV and VACV orthologs could distinguish prior mpox infection from vaccinia vaccination. Overall, these data highlight the use of multi-antigen panels in challenging scenarios for serological testing such as the cross-reactivity presented by orthopoxviruses.

## Introduction

Monkeypox virus (MPXV) is an enveloped, double-stranded DNA virus that belongs to the *Orthopoxvirus* genus of the *Poxviridae* family, and is the causative agent of mpox (1). In 2022, the Food and Drug Administration (FDA) and European Medicines Agency (EMA) approved the use of tecovirimat for treatment of mpox, though the drug has since failed to show clinical efficacy in three Phase III randomized control trials (2, 3). A vaccine – JYNNEOS or IMVANEX, a third-generation modified live-attenuated vaccinia virus (VACV) – has also been approved by both the FDA and EMA for the prevention of mpox and as post-exposure prophylaxis in adults aged 18 years and older (4, 5).

As of August 31, 2025, the global outbreak of mpox has resulted in more than 150,000 cases and 424 deaths (6). Diagnosis typically relies on PCR-based assays (7), though serological testing could aid in identifying asymptomatic infections, estimating population immunity, defining correlates of protection, and discriminating people with prior mpox from VACV-vaccinated individuals. Currently, no FDA-authorized serology test exists for mpox diagnosis or immunity, and only three mpox serology tests are currently being offered clinically in the United States – ELISAs for orthopoxvirus IgG or IgM and a microsphere-based immunoassay for orthopoxvirus IgG antibody detection (8, 9). High sequence similarity (94–98%) between MPXV and VACV surface proteins complicates serological discrimination due to cross-reactivity (10). In 2008, adsorption-based ELISA or Western blot was shown to distinguish between MPXV-infected versus VACV-vaccinated individuals, albeit in small populations (11, 12). Single-analyte ELISA or whole-virion serological tests have not been able to adequately separate these populations, though efforts to identify discriminatory antigens are ongoing (13–15).

Multi-antigen serological tests may have a greater ability to discriminate among these populations. ELISA testing separately conducted with 24 MPXV and 3 VACV antigens suggested MPXV A27L as a marker of MPXV infection and MPXV M1R (VACV L1R) as an indicator of JYNNEOS vaccination (16). A multiplexed microsphere immunoassay consisting of two MPXV-specific peptides and five cross-reactive OPV antigens demonstrated > 90% sensitivity and specificity in detecting people with prior mpox in vaccinated populations (17). Likewise, in a Luminex-based assay incorporating nine MPXV antigens and three VACV antigens, a panel of eight of these antigens collectively distinguished people with prior mpox or VACV-vaccinated individuals from unexposed controls with 98% sensitivity and 95% specificity. Among these positives, MPXV A27L was then used to distinguish MPXV-infected from VACV-vaccinated with sensitivity of 88% and specificity of 97% (14). However, neither of these assays are commercially available for clinical labs to run in-house.

The main commercially available multi-antigen panel for MPXV binding serology is the Meso Scale Discovery (MSD) V-PLEX Orthopoxvirus Panel 1 (IgG) Kit. Prior evaluations of the kit have shown that MPXV M1/VACV L1 antibody levels were higher on average in IMVANEX-vaccinated individuals compared to unexposed controls, consistent with previous findings (15, 18). Furthermore, MPXV M1/VACV L1 demonstrated greater sensitivity and specificity in distinguishing sera from vaccinated individuals from unexposed controls than in differentiating sera from people with prior mpox from unexposed controls. A similar trend was observed for MPXV A29L/VACV A27L, but in the opposite direction, showing improved performance in identifying people with prior mpox relative to unexposed controls (18). Among two MSD studies, MPXV E8L, performed best (or tied) to discriminate exposed (vaccinated or MPXV-infected) from unexposed (13, 18). Additionally, MSD antibody level results have been compared to MPXV or VACV neutralizing activity, but were not found to be strongly correlated (19). Here, we established the performance characteristics of the MSD V-PLEX Orthopoxvirus Panel 1 (IgG) Kit, including comparison to a MVA neutralizing antibody test, using sera taken from people with prior mpox, JYNNEOS vaccine recipients, a pediatric control cohort, and otherwise healthy adults under 40 years of age screened for anti-rubella virus IgG antibodies.

## Materials and Methods

### Specimens and Cohorts

Several cohorts of specimens were acquired for this study, and their demographic information is summarized in Table 1.

**Table 1:** Summary of demographic characteristics for each cohort.

|  | **Pediatric**<br>**(n = 68)** | **Rubella IgG<sup>+</sup>**<br>**(n = 111)** | **Vaccine**<br>**cohort A**<br>**(n = 22)** | **Vaccine**<br>**cohort B**<br>**(n = 30)** | **MPXV**<br>**2-6 mo**<br>**(n = 9)** | **MPXV**<br>**10 mo**<br>**(n = 17<sup>2</sup>)** |
| --- | --- | --- | --- | --- | --- | --- |
| **Male, n (%)** | 35 (51%) | 14 (13%) | 17 (77%) | 30 (100%) | 9 (100%) | 17 (100%) |
| **Female, n (%)** | 33 (49%) | 97 (87%) | 5 (23%) | 0 (0%) | 0 (0%) | 0 (0%) |
| **Age, mean (range)** | 9 (4 - 15) | 32 (19 - 42) | 37 (20 - 75) | 36 (20 - 65) | 36 (31 - 42) | 32 (20 - 49) |
| **HIV positive, n (%)** | NA | NA | 1 (5%) | 0 (0%) | 7 (78%) | 0 (0%) |
| **Born < 1972, n(%)** | 0 (0%) | 0 (0%) | 4 (18%) | 2 (7%) | 0 (0%) | 0 (0%) |
| **history of vaccination,**<br>**n (%)** | 0 (0%) | 0 (0%) | 6 (27%) | 0 (0%) | 0 (0%) | 0 (0%) |
| **Months post-MPXV,**<br>**mean (range)** | NA | NA | NA | NA | 4.2 (1.9 - 5.6) | 10.3 (7.9 - 12.4) |
| **Months post-vaccine,**<br>**mean (range)** | NA | NA | multiple<br>collections<sup>1</sup> | 9.1 (5.5 - 10.9) | NA | NA |

Abbreviations: months (mo)
<sup>1</sup>specimens were collected at baseline, 1 mo after first dose, and 2 mo after first dose
<sup>2</sup>One individual was sampled twice, once at 9.6 months and once at 11.6 months post MPXV infection, so total number of specimens analyzed was 18.

Two cohorts presumed to be unexposed to orthopoxviruses were included as negative controls for assay specificity and threshold determination. The first consisted of 111 deidentified remnant serum samples from adults born after 1981 with detectable anti-rubella IgG (Rubella IgG⁺ cohort) on the FDA-approved DiaSorin LIAISON Rubella IgG Assay, obtained from the University of Washington (UW) Virology Laboratory. The second included 68 deidentified remnant serum samples from pediatric patients undergoing allergen testing at Seattle Children’s Hospital (Pediatric cohort).

Two cohorts were acquired to evaluate responses associated with JYNNEOS smallpox vaccination. The longitudinal vaccine cohort (Vaccine A cohort) comprised 22 individuals with a complete set of serum specimens collected at baseline (Vaccine A - 0 month), 1 month after the first dose (Vaccine A - 1 month), and 1 month after the second dose (Vaccine A - 2 month), provided by the UW Virology Research Clinic (VRC). The cross-sectional vaccine cohort (Vaccine B) included 30 individuals sampled at an average of 9.1 months post-vaccination, provided by the Sexual Health Clinic (SHC) at Public Health-Seattle & King County (PHSKC).

Lastly, MPXV responses were assessed in two cohorts. The first cohort (MPXV – 2–6 month) consisted of nine individuals sampled at an average of 4.2 months post-infection, provided by the UW VRC. The second cohort (MPXV – 10 month) included 17 individuals sampled at an average of 10.3 months post-infection, provided by the SHC at PHSKC. One individual in the MPXV – 10-month cohort was sampled twice (at 9.6 and 11.6 months), resulting in a total of 18 specimens. All specimen use was approved by the University of Washington or Seattle Children’s Hospital Institutional Review Board with a consent waiver for control cohorts of remnant specimens and with written informed consent from the VRC and SHC/PHSKC cohorts.

### Production of MVA Virus

Vaccinia virus Modified Vaccinia Ankara strain (MVA) (BEI, NR-727) was propagated in BHK-21 cells (ATCC, CCL-10) at a multiplicity of infection (MOI) of 0.7. Cultures were harvested 72 hours post-infection, when approximately 80% of cells exhibited cytopathic effects, by scraping cells into the culture supernatant and transfer of the entire cell suspension to a 15 mL conical on ice. The cell suspension was centrifuged at 1,200 × g for 10 minutes at 4 °C to separate cell debris, and the resulting supernatant (S0) was transferred to a 15 mL conical tube and kept on ice. The cell pellet was resuspended in 1 mL of S0 and subjected to a freeze-thaw cycle by immersion in liquid nitrogen for 1–2 minutes, followed by rapid thawing at 37 °C. The resuspended freeze-thawed pellet was briefly vortexed and centrifuged again at 1,200 × g for 10 minutes at 4 °C. The supernatant from the freeze-thaw (FTS) was collected into a separate 15 mL conical tube on ice. This freeze-thaw procedure was repeated two additional times. Finally, all collected FTS supernatants were pooled and combined with the remaining S0.

### Vaccinia Virus Focus Reduction Neutralization Test

Flat-bottomed 96-well plates (Corning, 3585) were seeded with 10,000 VeroE6 cells (ATCC, CRL-1586) per well in 100 µL DMEM-10, which consisted of DMEM (Gibco, 10564-011), supplemented with 10% fetal bovine serum (Gibco, A38400-01) and 1% Penicillin–Streptomycin (Gibco, 15140-122) and incubated overnight at 37°C in 5% CO_2_.

Serum specimens were thawed and heat inactivated at 56°C for 30 minutes. 10.5 µL of sera were diluted into 94.5 µL of DMEM-10 in a polypropylene U-bottomed plate (Thermo Fisher, 267245) with one column left with only DMEM-10 to serve as a virus only control (VOC). A row at the top and bottom of the plates serve as media-only controls (MO). Sera were then serially three-fold diluted by transferring 35 µL 1:10 diluted sera into 70 µL complete DMEM-10, creating dilutions ranging from 10-fold to 2,430-fold. A 50 µL aliquot of the expanded MVA P1 passage was sonicated on ice using a cup horn at 50% power with 2 second pulse on/2 sec pulse off for 1 minute and then diluted 1,500-fold in complete DMEM-10. Then 70 µL of diluted virus was added to 70 µL diluted sera and VOC wells, diluting sera a further 2-fold, for final range of 20-fold to 4,860-fold. The virus/serum mixture was incubated at 37°C for 1 hour, the media was aspirated from pre-plated VeroE6 cells, and 50 µL of the virus/serum mixes, VOC controls, and MO controls was added to VeroE6 cells in duplicate and incubated for 24 hours at 37 °C. Some specimens were initially diluted by combining 21 µL of sera and 84 µL of complete DMEM-10 (final dilution series 10-fold to 2,430-fold) or by combining 5.25 µL of sera and 99.75 µL of complete DMEM-10 (final dilution series 40-fold to 9,720-fold). The initial dilutions utilized in testing are shown in Table S11.

Plates were fixed by adding 25 µL of 4% formaldehyde/DPBS to the media and incubated for 60 minutes at room temperature. Plates were washed three times with 150 µL PBS to remove formaldehyde. Plates were then treated with rabbit polyclonal anti-vaccinia virus antibody (ViroStat, 8101) diluted 1000-fold in FFA stain (1x PBS, 1 mg/mL saponin, and 0.1% BSA) in a total volume of 50 µL per well. Plates were incubated at room temperature for 2 hours and then washed three times with 150 µL FFA wash buffer (0.05% Triton-X/PBS) by incubating plates with wash for 2 minutes then discarding wash buffer. Plates were then treated with goat anti-rabbit IgG (Bethyl Laboratories, A120-201P) diluted 7,000-fold in 1x FFA stain in a total volume of 50 µL per well. Plates were incubated at room temperature for 1 hour in the dark and then washed with 150 µL FFA wash buffer as described above. Plates were stained with 50 µL 0.22 µm-filtered TrueBlue (SeraCare, 5510-0050) for exactly 15 minutes, then washed twice with 150 µL Milli-Q water. Plates were dried then counted with CTL BioSpot 7.0.38.7 imager.

The MVA neutralization assay was qualified. Briefly, precision was estimated and found acceptable (geometric standard deviation of the 50% neutralizing titer (ND50) < 1.42) by repeat testing specimens M33, V47, M48, M50, M8 in duplicate over three days (data not shown). The CBER Vaccinia IgG Reference Standard, Lot 1, yielded an average ND50 of 1,551, equivalent to 41.3 µg/mL, consistent with previously reported values of 24–49 µg/mL (20). The lower limit of quantification (LLoQ) of the MVA neutralization was defined by the lowest dilution tested.

### Meso Scale Discovery V-PLEX Multispot Orthopoxvirus Serology Assay

MPXV antibody titers were measured using the MSD V-PLEX Orthopoxvirus Panel 1 (IgG) Kit (MSD, K15688U-2), an electrochemiluminescence assay that quantitatively detects IgG antibodies against five MPXV viral proteins (A29L, E8L, M1R, A35R, and B6R) and their corresponding VACV homologs (A27L, D8L, L1R, A33R, and B5R) in human serum samples. Assay plates were first blocked by adding 150 µL of MSD Blocker A to each well, followed by incubation for at least 30 minutes with orbital shaking at 700 rpm (JitterBug, Model 270400). During the blocking step, serum specimens were diluted between 1:500 and 1:400,000 in MSD Diluent-100 (MSD, R50AA-2) in 1 mL polypropylene deep-well plates (USA scientific, 1896-1110). A calibrator series was prepared from a 10X calibrator stock (MSD, C0686-2) by first diluting 10-fold, then serially 4-fold over seven dilutions (CAL-01 through CAL-07). A blank control (CAL-08) was prepared using Diluent-100 alone. Following blocking with Blocker A, the assay plates were washed three times with 300 µL/well of 1X MSD Wash Buffer using an Agilent BioTek plate washer (Model 50TS9V-SN). Following the washing steps, 50 µL of diluted samples, Orthopoxvirus Serology Controls 1 through 3 (MSD, C4686-1), and calibrators were added to the assay plate. Plates were incubated for 2 hours at room temperature with orbital shaking at 700 rpm. After incubation, the plates were washed again as described above. Next, 50 µL of SULFO-TAG Anti-Human IgG Antibody (MSD, D21ADF-3) was added to each well, followed by incubation for 1 hour at room temperature with shaking at 700 rpm. After the antibody incubation, the plates were washed again as described above, and 150 µL/well of MSD GOLD Read Buffer B was added. The plates were read using the MESO QuickPlex SW 120MM instrument.

A detailed description of the MSD assay validation is provided in the Supplementary material. For precision testing, serum specimens V4, M31, V37, V54, and M60 (selected from the Vaccine B and MPXV–10-month cohorts) were thawed and single-use aliquots prepared for analysis. In addition to the calibrators, two additional specimens were used to assess linearity, M32 (from MPXV-10-month cohort) and Reference Vaccinia Immune Globulin (VIGIG), lot 1 from CBER/FDA (VIGIG). M32 was initially diluted 5-fold and VIGIG diluted 50-fold to bring the materials near the linear range of the assay. For testing, pre-diluted linearity specimens were diluted and tested following the same procedure as the calibrators. Table S1 summarizes the limit of detection (LoD), lower limit of quantification (LLoQ), and upper limit of quantification (ULoQ) for each antigen.

### Analysis and statistical methods

Antibody levels and MVA neutralization titers were log-transformed prior to analysis. For the MVA neutralization assay, the ND50 was estimated by four-parameter logistic modeling using the dr4PL R package (version 2.0.0) (21). Antibody levels in AU/mL were determined using the MSD Discovery Workbench Desktop Analysis software (version 4.0). ND50 and MSD estimates that fell below the LLoQ were retained as point estimates derived from curve fitting, rather than being imputed (e.g., as ½ LLoQ). One specimen, P61, had an undetected result for VACV B5R, and was set to an AU/mL of ½ LoD. Pearson’s correlation coefficients were calculated to assess relationships between antibody levels across ortholog pairs, while Spearman’s correlation coefficients were used to evaluate associations between antibody levels and MVA neutralization titers. Both correlation coefficients and associated p-values were computed using the cor.test function from the base stats package in R (version 4.5.1). Group comparisons of antibody levels, log₂-transformed ortholog pair ratios, and MVA neutralization titers were performed using two-sided t-tests without assuming equal variance. To account for multiple comparisons, p-values were adjusted using Bonferroni correction. The rstatix R package (version 0.7.2) was used to perform t-tests and apply p-value corrections (22). Receiver operating characteristic (ROC) curve analysis was conducted using the pROC R package (version 1.19.0.1) (23). For the ROC analysis baseline sera collected in the longitudinal vaccine group were included in the exposed group if they had a history of vaccination (6 of 22), otherwise, they were allocated to the negative group. All analysis code and figure generation scripts are available at https://github.com/greninger-lab/MPXV_MSD_validation_paper.git. The complete dataset is available in Table S11.

## Results

### Validation of MSD Orthopoxvirus antibody titer assay

The MSD V-PLEX Orthopoxvirus Panel 1 (IgG) assay quantifies antibody levels (in units of AU/mL) against five MPXV antigens (A29L, A35R, B6R, E8L, M1R) and their VACV orthologs (A27L, A33R, B5R, D8L, L1R). These orthologs are highly conserved (> 94% amino acid identity) and presumed to share functional properties (Figure 1A). VACV A33R and B5R are transmembrane proteins associated with the enveloped virion (EV) (24–26), while VACV D8L and VACV L1R are intracellular transmembrane proteins associated with the mature virion (MV) (27, 28) (Figure 1B). VACV A27L is peripherally associated with MV via non-covalent interactions (29).

**Figure 1:**
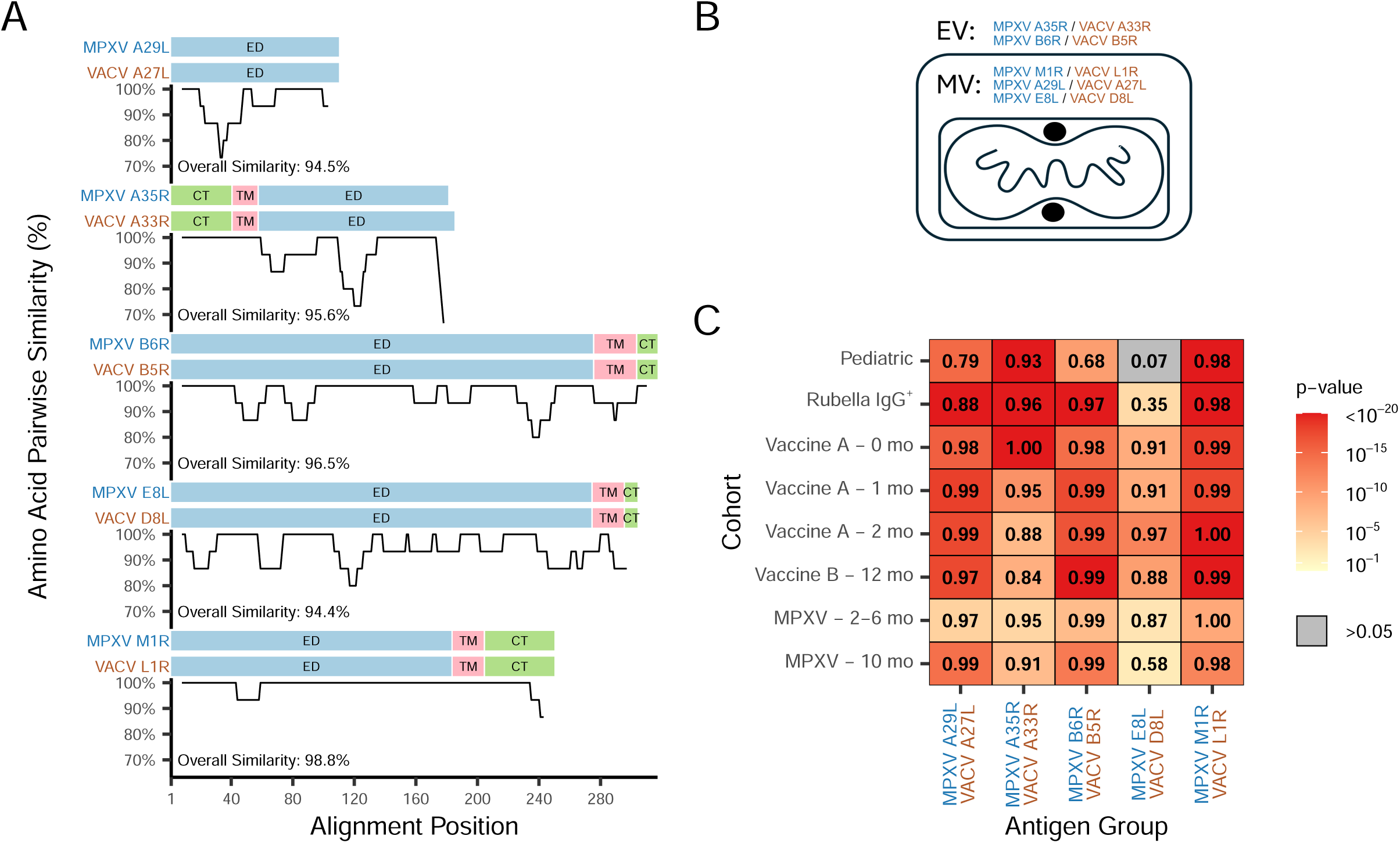
Responses to paired MPXV and VACV antigen orthologs are highly correlated in each cohort. (A) Paired MPXV and VACV antigen orthologs in the MSD orthopoxvirus antibody assay are highly conserved. Amino acid similarity is depicted for each ortholog pair using a 15 amino acid sliding window. Endodomain/cytoplasmic tail (CT), transmembrane (TM), and ectodomain (ED) regions are shown above each similarity graph. (B) Orthologs selected for the MSD assay are found either on the intracellular mature virion (MV) or extracellular virion (EV). (C) Pearson correlations of the antibody level for each paired MPXV and VACV ortholog are shown for each of the specimen cohorts examined. To determine if a correlation is significantly different from zero (i.e. no correlation), a t-test was performed and the heatmap is colored by the associated p-value of this test, as depicted in the legend key. The P61 VACV B5R result was omitted from the plot, since the antibody level was undetected.

Our validation of the MSD assay, following Good Clinical Laboratory Practices (GCLP) guidelines (Supplemental Material), confirmed acceptable performance for the calibrators and calibrator model (calibrator regression fits R^2^ > 0.9; Figure S1-S3 and Table S2), linearity (slope 0.9–1.0, R² > 0.9, intercept < 0.2 AU/mL; Figure S4), precision (within-lab geometric coefficient of variation (GCV) < 37%; Figure S5-S6), accuracy (serology control 1 and 2 %RE within ±30% and control 3 < 0.2 AU/mL; Figure S7), and freeze-thaw robustness (0.5–1.5-fold variation over three freeze-thaws; Figure S8). Minor deviations (e.g. elevated CAL-08 signal for M1R/L1R) did not affect overall performance (Figure S2). The assay was deemed suitable for quantitative serology and subsequent cohort testing.

### Anti-orthopoxvirus IgG antibody levels are elevated in sera from vaccinated and MPXV-infected individuals

We assessed serological responses to MPXV infection and vaccination on the MSD assay across six cohorts, including two negative cohorts, two JYNNEOS-vaccinated cohorts, and two MPXV-infected cohorts (see Methods). Antibody responses to orthologs were strongly correlated across exposed groups (⍴ > 0.8 for most comparison; Figure 1C), consistent with sequence conservation. MPXV E8L and VACV D8L exhibited weaker correlations, especially among presumed negative cohorts and MPXV-infected individuals due to distinct responses to the ortholog pairs (Figure S9).

We next compared antibody responses across cohorts for each ortholog pair (Figure 2) and evaluated statistical significance of observed differences (Table S3; Figure S10).

**Figure 2.**
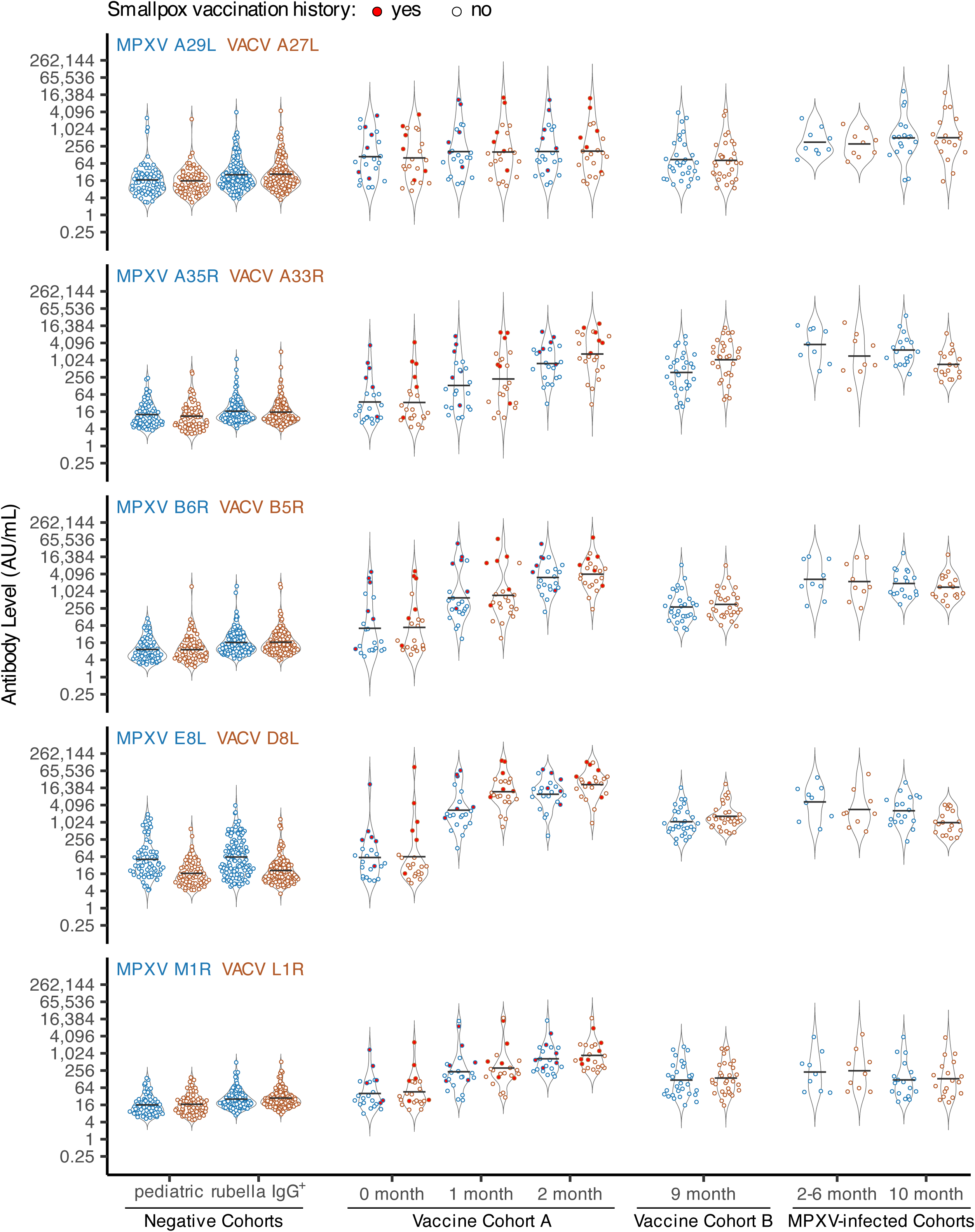
Anti-orthopoxvirus IgG antibody levels are elevated in vaccinated or MPXV-infected individuals. Eight groups of specimens from six cohorts (x-axis) were tested using the MSD assay, and Signal values were converted to AU/mL. Responses to MPXV orthologs are shown in blue and VACV orthologs in brown. The ortholog pair for each plot is indicated in the upper left corner. Black lines represent group geometric mean titers (GMT), and untrimmed violin plots show the distribution behind individual data points. The points corresponding to individuals with a known history of smallpox vaccination are filled in with red.

Negative cohort geometric mean titers (GMTs) were generally low, though antibody levels overlapped with the exposed cohorts. Adult rubella IgG^+^ specimens tended to have slightly higher GMTs compared to the pediatric cohort (e.g., VACV L1R: Pediatric 16.7 AU/mL vs. Rubella IgG+ 27.7 AU/mL; p-value < 0.05). Within the negative cohorts, GMTs were similar across ortholog pairs, except for MPXV E8L/VACV D8L (e.g., Pediatric: MPXV E8L 51.4 AU/mL vs. VACV D8L 16.7 AU/mL; p-value < 0.05).

Baseline GMTs in the Vaccine A cohort trended higher relative to the negative cohorts. When individuals with a known history of smallpox vaccination were removed (see red-filled points, Figure 2), Vaccine A baseline GMTs were similar to the negative cohorts, except for MPXV A29L/VACV A27L (data not shown). Post-vaccination GMTs for Vaccine A and B cohorts were generally higher than negatives and Vaccine A baseline (0 month), except for MPXV A29L/VACV A27L, which had modest increases which were significant only when compared to the Pediatric cohort (e.g. MPXV A29L: Pediatric vs. Vaccine A – 1 month, 16.9 vs. 166.4 AU/mL, p-value < 0.05; MPXV A29L: Vaccine cohort A – 0 month vs. Vaccine cohort A – 1 month, 110.0 vs. 166.4 AU/mL, p-value > 0.05). Within the Vaccine A cohort, GMTs increased from baseline to 2 months post-vaccination (e.g. MPXV E8L: Vaccine A, 1 month vs. 2 months, 2,710.5 vs. 9,849.5 AU/mL, p-value > 0.05), except for MPXV A29L/VACV A27L. In Vaccine B, GMTs at 10 months post-vaccination were significantly lower than those in Vaccine A at 2 months (e.g. MPXV B6R: Vaccine B – 9 months vs. Vaccine A – 2 months, 285.0 vs. 3,099.1 AU/mL; p-value < 0.05), consistent with waning antibody responses.

Across all ortholog pairs, MPXV-infected cohorts had significantly higher GMTs than negatives (p-value < 0.05, with limited exceptions within M1R/L1R). Unlike in the vaccinated cohorts, MPXV-infected cohorts showed significantly higher MPXV A29L/VACV A27L titers relative to negatives (p-value < 0.05).

### Antibody level ratios between orthologs reveal distinct responses in infected vs. vaccinated cohorts

To investigate differences in antibody responses between MPXV and VACV orthologs, we calculated the ratio of MPXV to VACV antibody responses for each ortholog pair and compared mean ratios between cohorts using two-sided t-tests (Figure 3 and Table S4). For MPXV A29L/VACV A27L and MPXV M1R/VACV L1R, mean ratios did not differ significantly between exposed groups. In contrast, significant differences in MPXV/VACV response ratios were observed for MPXV A35R/VACV A33R, MPXV B6R/VACV B5R, and MPXV E8L/VACV D8L, with MPXV-infected cohorts consistently showed higher MPXV/VACV ratios, while vaccinated cohorts showed lower MPXV/VACV ratios. For example, the MPXV A35R/VACV A33R mean ratio was significantly elevated in MPXV-infected cohort at 2–6 months post-infection compared to vaccinated cohorts at 1, 2, and 9 months (2.55 vs. 0.59, 0.46, and 0.36, respectively; p-values < 0.01). The decline in MPXV A35R/VACV A33R ratio in vaccinated cohorts suggests a shift toward relatively higher VACV A33R responses longitudinally. Conversely, the MPXV E8L/VACV D8L ratio increased over time in vaccinated cohorts at 1, 2, and 9 months (0.23, 0.46, and 0.65, respectively), indicating a relative increase in MPXV E8L reactivity. Finally, the Pediatric and Rubella IgG+ cohorts exhibited elevated MPXV E8L/VACV D8L ratios (3.09 and 2.96, respectively), driven mostly by higher MPXV E8L responses, potentially reflecting non-specific binding to this antigen.

**Figure 3:**
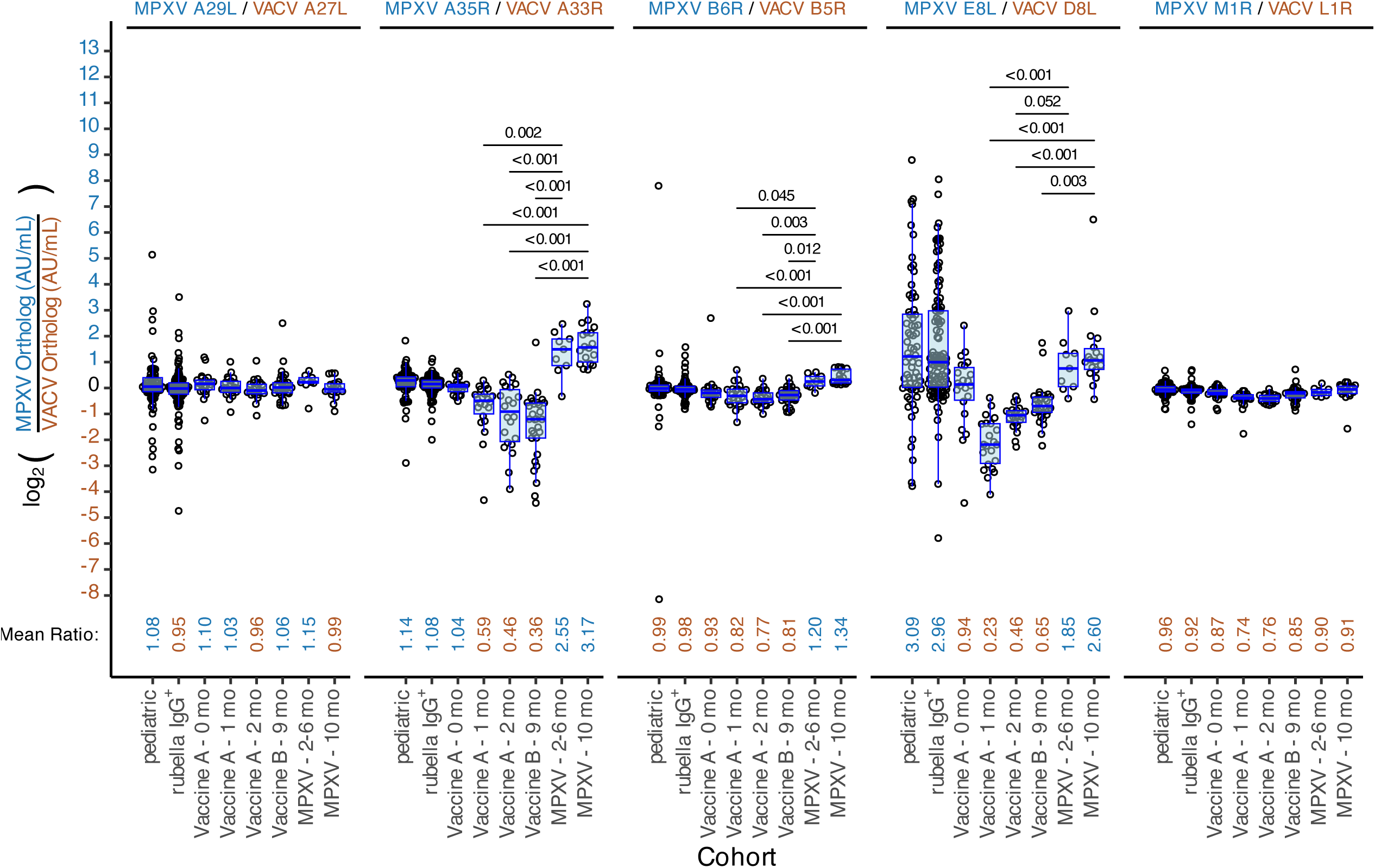
The response ratio between MPXV and VACV orthologs were significantly different for several ortholog antigen pairs. For each result, the ratio of the AU/mL result of the MPXV ortholog over the VACV ortholog was calculated, log_2_-transformed, and plotted for each ortholog pair and cohort. The y-axis labels have been colored such that positive-log-fold changes are blue (MPXV > VACV ortholog response) and negative-log-fold changes are brown (MPXV < VACV ortholog response). Overlaid is a box plot with the central line marking the median value, the hinges indicating the first and third quartile or interquartile range (IQR), the whiskers extending to the highest or lowest value within 1.5 times the IQR range. The mean ratio for each cohort was calculated and displayed at the bottom of each graph. Mean ratio values are colored such that responses that are stronger to the MPXV ortholog are colored blue and responses that are stronger to the VACV ortholog are colored brown. Utilizing the mean ratio values, we performed a t-test comparing ratios between cohorts using a Bonferroni correction for multiple comparisons. Significant t-test (p-value < 0.05) are shown in the upper part of each graph, only for the comparisons between the MPXV-infected and MVA-BN vaccinated, see Table S4 for all comparisons. The horizontal black bar indicates the groups being compared for a given p-value.

### Sera from vaccinated or MPXV infected individuals were neutralizing against MVA

To assess the correlation between anti-orthopoxvirus antibody levels determined by the MSD assay and functional neutralizing activity, we performed a focus reduction neutralization test (FRNT) using MVA to quantify ND50 titers in all cohorts (Figure 4A). Two-sided t-tests were used to compare ND50 titers between groups. Overall, ND50 titers were low across cohorts, consistent with previous reports of orthopoxvirus neutralization assays that lack complement (30). The negative and Vaccine A baseline cohorts exhibited minimal neutralizing activity (ND50 < 20), with a few exceptions: one Rubella IgG+ specimen (RU6, ND50 = 26.3) and two Vaccine A baseline specimens from individuals with a history of smallpox vaccination (VRC30, ND50 = 162.6; VRC26, ND50 = 30.7; Table S11). The higher ND50 GMT of the Vaccine A baseline group, relative to the negative cohorts, in part reflects prior smallpox vaccination in some individuals (Figure 4A, red colored points). Following vaccination, Vaccine A participants showed a marked increase in ND50 GMTs: a 6.6-fold rise from baseline to 1 month (ND50 GMT 5.3 to 35.2), and a further 2.1-fold increase from 1 to 2 months (ND50 GMT 35.2 to 74.4). Relative to Vaccine A cohort participants at 2 months post-vaccination, Vaccine B cohort participants (9 months post-vaccination) had 3.1-fold lower ND50 GMTs (ND50 GMT 24.1 vs. 74.4), again consistent with waning neutralizing titers. Among MPXV-infected individuals, ND50 GMTs were comparable between the 2–6 month and 10-month post-infection groups (ND50 GMT 41.2 vs. 30.1), and similar to titers observed in Vaccine A (1 month) and Vaccine B (9 months) cohorts. The highest neutralizing responses were observed in Vaccine A cohort individuals with prior smallpox vaccination.

**Figure 4:**
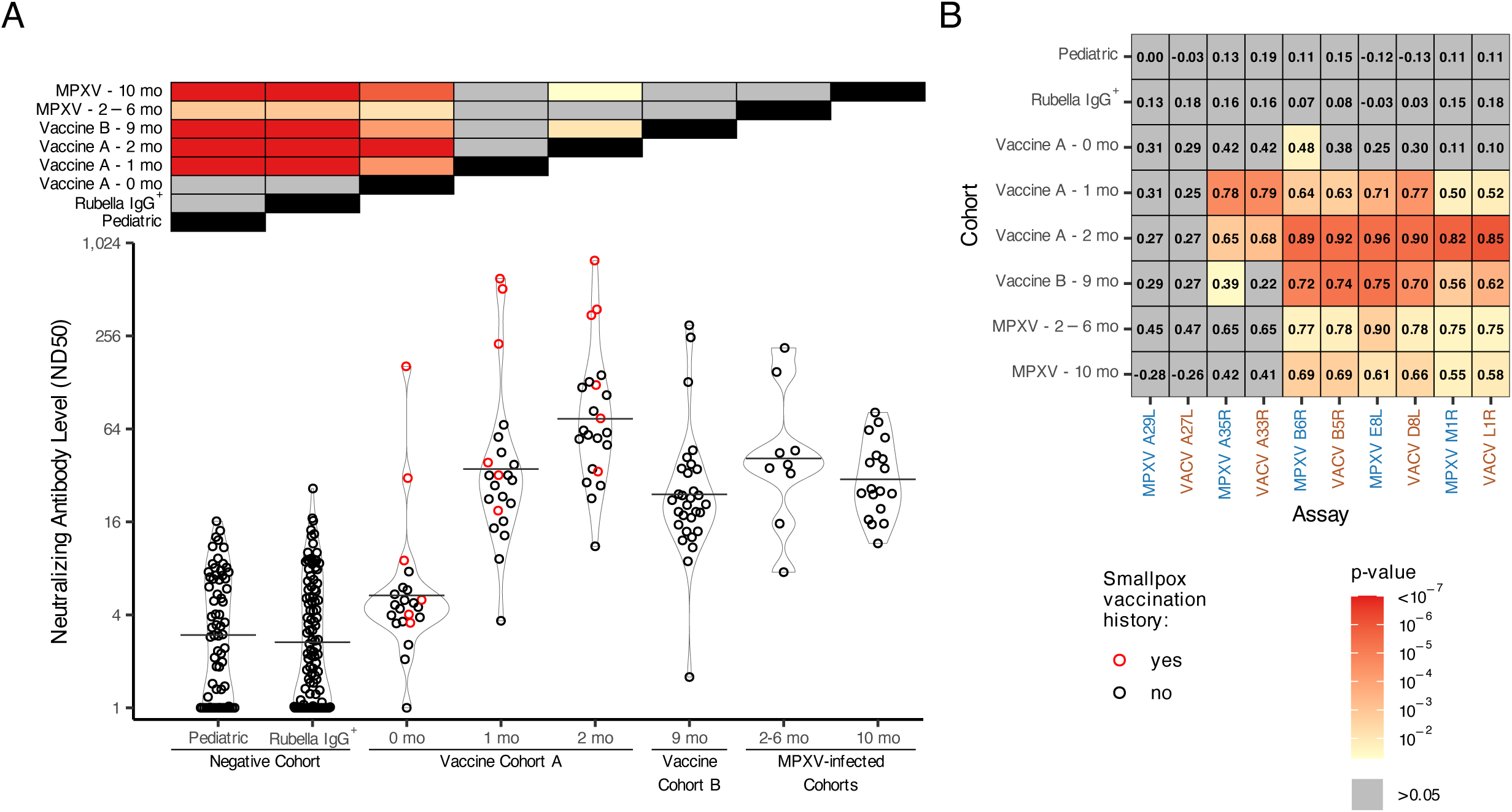
Sera from vaccinated (post-baseline) or MPXV-infected exhibited low-level neutralization of Vaccinia MVA, which was generally correlated with anti-orthopoxvirus antibody level. (A) The ND50 value against MVA was determined for all the sera tested in the MSD orthopoxvirus antibody titer assay and plotted by cohort. Prior smallpox vaccination history is indicated by red filled points. The black horizontal lines correspond to each cohorts GMT. Two-sided t-tests were used to compare ND50 titers between groups, and Bonferroni-adjusted p-values were visualized in a heatmap along the top of the figure with yellow-to-red shading indicating statistical significance (p-value < 0.05). (B) Sperman’s correlations were calculated between antibody titer results for each antigen and the MVA neutralization ND50 results within each cohort separately. To determine if a correlation is significantly different from zero (i.e. no correlation), a t-test was performed and the fill value in each tile corresponds to the p-value of this test as shown in the legend key.

We next compared MSD-derived antibody levels with MVA FRNT ND50 values across all antigens and cohorts (Figure 4B and S11). In the negative cohorts (Pediatric and Rubella IgG+) and the Vaccine A baseline group (0 months), binding titers and neutralization were poorly correlated (Spearman ⍴ < 0.5), with individual responses randomly distributed between antibody level and ND50 (Figure 4B and S11). A similar pattern was observed in the Vaccine A baseline cohort, except for 2–3 individuals with prior smallpox vaccination, who showed stronger correlation between ND50 and binding titers (Figure S11, red-filled points). In the Vaccine A cohort, the correlation between antibody titer and MVA neutralization strengthened over time following vaccination for all antigens except MPXV A29L/VACV A27L and MPXV A35R/VACV A33R. Neutralizing titers post-vaccination were most strongly correlated with antibody binding levels to MPXV B6R/VACV B5R, MPXV E8L/VACV D8L, and MPXV M1R/VACV L1R. Interestingly, the correlation between MPXV A35R/VACV A33R and MVA neutralization appeared strongest 1 month post-vaccination but declined at 2 months post-vaccination. MPXV-infected individuals showed a similar antigen-specific correlation pattern to post-baseline vaccinated cohorts.

### Diagnostic performance of the MSD assay using single antigens or ortholog pair response ratios

To evaluate if antibody levels for individual antigens can distinguish people with prior mpox from negative controls (Rubella IgG+ and Pediatric) or vaccinated from negative controls, we performed receiver operating characteristic (ROC) analysis. Starting with MPXV-infected verses negative controls, area under the curve (AUC) values ranged from 0.892 to 0.998, with MPXV A35R and MPXV B6R showing the highest performance (Figure S12 and Table S5). Youden index thresholds determined for MPXV A35R and MPXV B6R were 295.6 AU/mL and 252.1 AU/mL, respectively. At these thresholds, each antigen had a specificity of 1.000 (95% CI 1.000 – 1.000) and sensitivity of 0.990 (95% CI 0.974 - 1.000) for MPXV A35R and 0.979 (95% CI 0.959 – 0.995) for MPXV B6R. The VACV orthologs of these antigens, VACV A33R and VACV B5R, showed similar performance with sensitivity/specificity > 0.95. MPXV M1R and VACV L1R, exhibited the lowest AUCs (0.895 and 0.891, respectively) and specificity/sensitivity < 0.9. In the comparison of vaccinated verses negative controls, AUC values ranged from 0.785 to 0.991, with VACV D8L showing the highest performance (Figure S12 and Table S6). VACV D8L achieved a specificity of 0.988 (95% CI 0.963 – 1.000) and sensitivity of 0.969 (95% CI 0.949 –0.995) at 233.1 AU/mL. Its MPXV ortholog, E8L, performed less well (AUC 0.946), likely due to broader antibody level distributions in the negative cohorts (Figure 2 and Figure S13). MPXV A29L and VACV A27L, exhibited the lowest AUCs (0.792 and 0.785, respectively) and specificity/sensitivity < 0.9.

We also evaluated performance of the assay to distinguish exposed (MPXV-infected and vaccinated combined) from negative controls (Rubella IgG^+^ and pediatric). AUC values ranged from 0.818 to 0.991, with VACV D8L showing the highest performance (Figure 5A and Table S7). VACV D8L achieved a specificity of 0.991 (95% CI 0.972 – 1.000) and sensitivity of 0.969 (95% CI 0.944 – 0.990) at 233.1 AU/mL (Table S7). As with the vaccinated vs. negative control comparison, the MPXV E8L ortholog performed less well (AUC 0.953).

**Figure 5:**
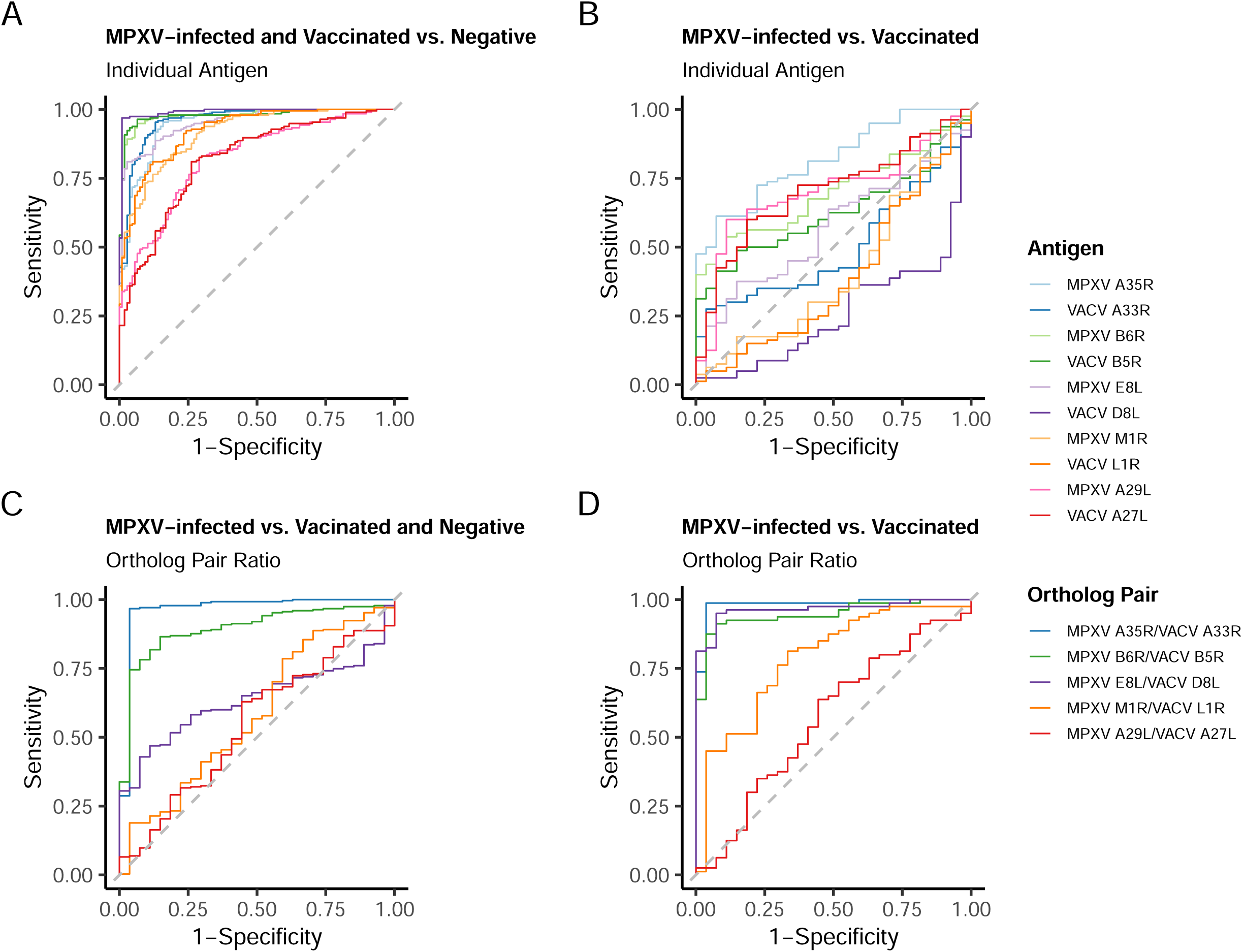
Receiver operator characteristic (ROC) curves utilizing either the anti-orthopoxvirus antibody level results or the ratio of antibody level to ortholog pairs. MPXV-infected refer to all sera from known MPXV infected individuals, Vaccinated refers to all sera from known vaccinated individuals, and Negative refer to sera from the rubella and pediatric negative cohorts combined. Baseline sera collected prior to vaccination (Vaccine A – 0 month) were included in the Vaccinated group if they had a history of smallpox vaccination, otherwise they were categorized as negative (6 out of 22 individuals had previous history of vaccination). (A) ROC curves for all individual antigens generated by comparing all known MPXV-infected or Vaccinated (n = 107) to all Negative (n = 195). (B) ROC curves for all individual antigens generated by comparing all known MPXV-infected (n = 27) to all known Vaccinated (n = 80). (C) ROC curves for all antigen ortholog pairs generated by comparing antigen ortholog pair ratios from known MPXV-infected (n = 27) to Vaccinated and all Negative (n = 275). (D) ROC curves for all antigen ortholog pairs generated by comparing antigen ortholog pair ratios from known MPXV-infected (n = 27) to known Vaccinated (n = 80). Summary statistics from this analysis can be found Tables S7-S10.

Next, ROC analysis was performed to evaluate performance of the assay to distinguish specimens from MPXV-infected from vaccinated individuals. Here lower AUCs (0.260–0.819) were observed, with MPXV A35R performing best (AUC 0.819; Figure 5B and Table S8). However, sensitivity was limited (0.613 at 621.1 AU/mL), reflecting the challenge of differentiating infection from vaccination using single-antigen thresholds.

Based on Youden index thresholds determined on differentiating specimens from exposed or negative individuals (Figure 5A and Table S7), 60% (41/68) of Pediatric and 64% (71/111) of Rubella IgG+ specimens exceeded the cutoff for at least one antigen (Figure S13). Two Rubella IgG+ specimens exceeded thresholds for all ten antigens and were MVA neutralizing (RU6, ND50 26.3; RU58 ND50 13.3; Figure S12), suggesting prior orthopoxvirus exposure or vaccination. Other specimens exceeded fewer thresholds and lacked neutralization, indicating possible cross-reactivity.

Since we identified that certain ortholog pairs showed differential responses between vaccinated and MPXV-infected individuals (Figure 3), we next assessed the discriminatory power of the ortholog pair ratios. We generated ROC curves comparing MPXV-infected specimens to vaccinated and negative cohorts (Figure 5C and Table S9). MPXV A35R/VACV A33R and MPXV B6R/VACV B5R ratios performed best, with AUCs of 0.965 and 0.894, respectively. For MPXV A35R/VACV A33R, the optimal threshold was a ratio of 1.6, yielding 0.963 specificity (95% CI: 0.889–1.000) and 0.967 sensitivity (95% CI: 0.945–0.989). MPXV E8L/VACV D8L had poor performance when negatives were included (AUC 0.639) but improved when negatives were excluded with an AUC 0.965, specificity 0.926 (95% CI 0.852 – 1.000), sensitivity of 0.950 (95% CI 0.812 - 1.000; Figure 5D and Table S10). In addition, when negatives were excluded, MPXV A35R/VACV A33R and MPXV B6R/VACV B5R ortholog ratios AUCs increased from 0.965 to 0.983 and 0.894 to 0.946, respectively.

## Discussion

Here we show results from validation and testing of a commercial MSD multiplex antibody assay, confirming it meets GCLP criteria. To assess clinical performance characteristics, sera was tested in the MSD assay from presumed unexposed individuals, vaccinated individuals, and individuals with prior mpox. Both vaccination and MPXV infection induced measurable antibody responses to all MPXV and VACV antigens in the MSD assay, but vaccination and infection cohorts differed in the magnitude and pattern of responses. Antibody levels correlated with MVA-neutralization for most ortholog pairs, further corroborating assay performance. Finally, ROC analysis demonstrated that while single antigens could distinguish exposed from unexposed individuals, ortholog response ratios more effectively differentiated infection from vaccination.

Testing of these cohorts revealed some expected patterns. Antibody levels to MPXV A29L/VACV A27L showed little change post-vaccination, consistent with poor induction by third-generation smallpox vaccines (Figure 2) (14, 18, 31–34). In contrast to these results, older attenuated vaccine strains have been observed to elicit stronger VACV A27L responses (33, 35). Review of published MVA and MVA-BN genome sequences indicate that the VACV A27L ORF is intact (GenBank: U94848 and DQ983238, respectively), suggesting that further attenuation of the third-generation vaccine strains may have impaired A27L immunogenicity. In addition, even when VACV A27L responses are induced by vaccination, they have not been found to constitute a major fraction of neutralization activity (35, 36). Although not a longitudinal sampling, at 9 months post-vaccination (Vaccine B), antibody GMTs to all antigens were generally lower than those observed at 2 months post vaccination (Vaccine A), aligning with reports of waning antibody levels post-vaccination (37, 38) (Figure 2). Among single antigens, VACV D8L, VACV B5R, MPXV B6R, VACV A33R, and MPXV A35R best distinguished individuals exposed by either natural MPXV infection or vaccination, consistent with prior MSD assay studies (13, 18). When focusing on MPXV-infected-only versus negative controls MPXV A35R and MPXV B6R performed best, and when focusing on vaccinated-only versus negative controls VACV D8L performed best. Custom MSD plates (39) and multiplex bead-based platforms (14, 17) that utilized an overlapping but distinct set of antigens, also generally identified these antigens as discriminatory, except MPXV A35R did not perform as well in the custom MSD assay (39). We also observed that MPXV M1R/VACV L1R antigens showed better sensitivity and specificity in distinguishing specimens from vaccinated individuals from negative controls compared to the MPXV-infected specimens, consistent with stronger MPXV M1R/VACV L1R responses in the vaccinated relative to the MPXV-infected (Figure 2). (16, 18). Likewise, the opposite dynamic was observed for the MPXV A29L/VACV A27L ortholog pair due to weaker responses among the vaccinated (Figure 2) (18). However, overall, MPXV M1R/VACV L1R and MPXV A29L/VACV A27L, consistent with other studies, were generally poor or inconsistent markers (13, 14, 17, 18, 39)

Although, antigens included in the MSD assay generally distinguished exposed from unexposed individuals, no single antigen separated MPXV-infected from vaccinated (Figure 5B). Prior strategies include targeting antigens absent in MVA-BN, such as MPXV A27L (14, 16), MPXV A26L (39), and B21R (7, 12, 17) or those with exposure-dependent responses (not ortholog dependent), such as MPXV A29L/VACV A27L (elevated in MPXV-infected) and MPXV M1R/VACV L1R (elevated in vaccinated)(14, 16, 18). Although the MSD assay includes these latter two sets of orthologs and we observed exposure-dependent responses (Figure 2), these orthologs did not appear to perform well in distinguishing MPXV-infected from vaccinated in our data (Figure 5B and Table S8). Differential ortholog ratios have been reported elsewhere (14, 16–18, 40) and one group combined MPXV B6R/VACV B5R ratio and MPXV A27L to distinguish MPXV-infected from vaccinated (39). In this study, MPXV A35R/VACV A33R, MPXV B6R/VACV B5R, and MPXV E8L/VACV D8L ratios could discriminate MPXV-infected from vaccinated, with the strongest predictor being MPXV A35R/VACV A33R (Figure 5C and Table S9). One limitation of this approach is differential responses may be less obvious when individuals are both vaccinated and MPXV-infected (17). Continued evolution of MPXV surface antigens in future outbreaks may also lead to changing responses between MPXV and VACV orthologs.

Since these antigens are involved in orthopoxvirus neutralization, we further validated the MSD assay by assessing whether antibody levels correlated with MVA neutralization. Significant positive correlations were observed in post-baseline vaccinated and MPXV-infected individuals for all antigens except MPXV A29L/VACV A27L (Figure 5). A prior study using the same assay reported weaker correlations with neutralization, likely due to differences in the neutralization target (MPXV Clade IIb B.1 or VACV WR instead of MVA) and unknown exposure histories, limiting direct comparison (19).

One challenge of validating orthopoxvirus serological assays is obtaining specimens known to be unexposed to orthopoxvirus either through natural infection or more often vaccination. Although the negative cohorts assembled here likely lacked smallpox vaccination since they were born after 1972 and had no known background of orthopoxvirus exposure, two Rubella IgG^+^ specimens (RU6 and RU58) showed elevated antibody levels and MVA-neutralization, suggesting prior vaccination or exposure, though unconfirmed. Pediatric specimens have a higher likelihood of not having orthopoxvirus exposure and have been used by others to establish threshold values in the MSD assay (18). However, pediatric specimens may not be fully representative of the responses observed in adults. In fact, in this study Rubella IgG^+^ GMTs within orthologs consistently trended higher compared to the Pediatric GMTs. Based on thresholds for distinguishing exposed vs. unexposed individuals (Table S7), there were many negative specimens that exceeded these thresholds for a subset of antigens (commonly 1-2 antigens, Figure S12). Cross-reactive responses are not uncommon in these assays (16, 17, 41). MPXV E8L appears to have significant background or cross-reactive responses in both our negative cohorts (Figure 2 and Figure S12). Requiring positivity for greater than one antigen may improve assay performance (14, 17, 18).

There are several limitations of this study. First, we did not have individuals that were both vaccinated and MPXV-infected, which might affect differential response to the MPXV/VACV ortholog pairs, as observed elsewhere (17). The lack of an international standard for antiserum to VACV or MPXV complicates commutability of this work, although the calibrators provided by MSD should allow for harmonization of results between labs utilizing the commercial MSD assay. Our MPXV-infected cohorts were relatively small (total 27 specimens), which may impact ROC analysis. We only assessed neutralization against the attenuated MVA strain and correlations between antibody level and neutralization might differ if we also tested against MPXV or non-attenuated VACV. The vaccinated and MPXV-infected cohorts were strongly skewed towards men (77%-100% male) relative to the Rubella IgG^+^ cohort (13% male) and the Pediatric cohort (51% male), though this adequately reflects the population most at risk of MPXV infection. Lastly, though we performed Youden index analysis to determine post-hoc thresholds for exploratory purposes, we lack a validation cohort to test whether these thresholds confer the expected sensitivity and specificity. Overall, we demonstrate that the MSD orthopoxvirus IgG assay meets GCLP-based validation standards and can reliably distinguish orthopoxvirus-exposed from unexposed individuals. Furthermore, differential antibody responses to specific MPXV/VACV ortholog pairs (particularly MPXV A35R/VACV A33R and MPXV B6R/VACV B5R) may be useful for discriminating JYNNEOS-vaccinated from MPXV-infected individuals. Antibody levels measured by the MSD assay were generally correlated with MVA neutralization activity, supporting its relevance as a surrogate marker of the immune response. However, limited discriminatory performance for the MPXV A29L/VACV A27L and MPXV M1R/VACV L1R antigen pairs, along with the elevated background observed for MPXV E8L, highlights areas for further assay refinement. Inclusion of additional antigens, particularly those with divergent immunogenicity between MPXV and VACV or those that are missing in the attenuated vaccine strains, could further improve the assays utility to identify prior exposure and differentiate between routes of exposure. Given waning global population immunity to orthopoxviruses, MPXV outbreaks are likely to persist, and robust serological assays will be critical for understanding MPXV infection prevalence, defining immune correlates, and guiding vaccine policy and booster strategies.

## Supporting information

SuppTables

Line-item testing data is available in Table S11.

## Acknowledgements

ALG reports contract testing to UW from Abbott, Cepheid, Novavax, Pfizer, Janssen and Hologic, research support from Gilead, and personal fees from Arisan Therapeutics, outside of the described work. This research was funded in part by a 2022 developmental grant from the University of Washington/Fred Hutch Center for AIDS Research, an NIH funded program under award number AI027757 which is supported by the following NIH Institutes and Centers: NIAID, NCI, NIMH, NIDA, NICHD, NHLBI, NIA, NIGMS, NIDDK. The content is solely the responsibility of the authors and does not necessarily represent the official views of the National Institutes of Health.

## Supplementary Material

### Validation

#### Analytical sensitivity

The manufacturer does not specify a fixed limit of detection (LoD) for the VPLEX Orthopoxvirus assay, and instead the LoD is calculated for each run of the assay. The default method for calculating the LoD in the MSD Discovery Workbench 4.0 software utilizes the CAL-08 Signal results as follows:

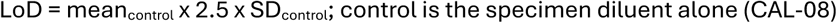

Once the LoD is calculated, in terms of Signal, it is converted to AU/mL utilizing the calibrator curve. To establish a fixed LoD, calibrators were run in duplicate over ten days (Figure S1A for results in Signal and S1B for results in AU/mL). Comparison of Signal results for each of the calibrators over time were distinct across all calibrators for antigens MPXV A29L, VACV A27L, MPXV A35R, VACV A33R, and VACV D8L. In contrast, antigens MPXV B6R, VACV B5R, MPXV E8L, MPXV M1R, and VACV L1R there was overlap in Signal between calibrators over time, especially the lower concentration calibrators and the blank (CAL-05 thru CAL-07 and CAL-08). Standardization to AU/mL largely resolves this overlap since it normalizes Signal dìerences between runs except for CAL-07 and CAL-08 for some antigens (MPXV M1R and VACV L1R). The LoD was determined for each antigen using the MSD Discovery Workbench software over the ten runs and the geometric mean of the LoD for each antigen ranged from 0.0031 – 0.0038 AU/mL. Thus, we set the LoD for each antigen at 0.004 AU/mL (Table S1)

For quality control an upper limit was set for CAL-08 of a Signal of 300 based on the range of values observed during our validation runs (Figure S2 – validation runs annotated with light blue bar). Notably, during validation, four out of ten runs exhibited higher CAL-08 Signal relative to the other runs for the MPXV M1R and VACV L1R antigens. For these antigens, the percent relative error (%RE) of the control materials provided by the manufacture were within +/- 30% for control 1 and 2 and < 0.2 AU/mL for control 3 and the geometric standard deviation of the control materials were all within 1.42. Thus, despite the slightly elevated CAL-08 Signal the assays were considered valid. During subsequent testing, several assays were flagged for having CAL-08 Signal exceeding a Signal of 300, especially for the MPXV M1R and VACV L1R antigens (Figure S2 between Oct 2024 and Jan 2025). Again, the manufactures controls performed as expected based on criteria described above. Plate images and data were provided to the manufacturer, and the manufacturer could not identify any artifacts (e.g. well drying) or manufacturing correlates that could explain the elevated CAL-08 Signal. All assays were performed by two operators using the same lot number of materials. To ensure sùicient dynamic range in Signal, the criteria was changed to ensure at least a 200-fold dìerence between CAL-01 and CAL-08 signals, rather than utilizing an absolute value for CAL-08 Signal. Based on this criterion, out of 75 assays, four assays failed for the M1R antigen and one assay failed for both M1R and L1R antigen (Figure S2, red-filled points).

#### Verification of Calibration model

The calibration model was verified by testing of the calibrators over ten days. The day-to-day percent relative error was within +/- 30% for all calibrators, except CAL-07 which was inconsistent from day-to-day for some antigens (data not shown). The average percent relative error observed over ten days of testing was within +/- 15% (Figure S3A). Finally, within-lab imprecision was within 15% geometric coèicient of variation (GCV) for all calibrators, except CAL-07 which was greater than GCV of 15% for a subset of the antigens (Figure S3B). Lastly, the four-parameter logistic regression (4PL) fits for each antigens calibration curve had R^2^ values > 0.9 (Table S2). Taken together, these data verify the validity of the calibration model and that CAL-07.

Based on the imprecision of the calibrators shown here the lower limit of quantification (LLoQ), was less than the LLoQ reported by the manufacturer (Table S1). The more conservative manufacturers estimates were selected as the preliminary LLoQ (see Table S1, LLoQ final column). The ULoQ was set to the mean back-calculated AU/mL value of the highest calibrator (CAL-01) (see Table S1, ULoQ final avg. calculated AU/mL of CAL-01).

#### Linearity

To assess linearity the results within the limits of quantification (LoQ) from testing of the assay calibrators over ten days were subjected to linear regression analysis (Figure S4A). For all antigens, the calibrators demonstrated strong linearity with R² values exceeding 0.9. Furthermore, for all antigens, the slopes of the linear regression were approximately one, and the y-intercepts were below 0.2 AU/mL, indicating the absence of systemic or proportional bias.

We also assessed linearity with strongly positive serum specimen (M32) that was previously tested in the V-PLEX Orthopoxvirus assay. The specimen was initially diluted 50-fold followed by seven 4-fold dilutions. Each dilution was measured in the assay and the back-calculated AU/mL values, not corrected for dilution, were subjected to linear regression (Figure S4B). Like the calibrators, the M32 dilution series was strongly linear with R² values exceeding 0.9. Furthermore, for all antigens, the slopes of the linear regression were approximately one, and the y-intercepts were below 0.2 AU/mL, indicating the absence of systemic or proportional bias. Lastly, the M32 dilution series results were highly parallel to the calibrator linear regression (Figure S4B, dotted line corresponds to the calibrator linear regression best fit), supporting the use of the calibrators and selected diluent in estimating AU/mL levels from serum specimens. Similar results were obtained with Reference Vaccinia Immune Globulin (VIGIG) pooled human serum from CBER/FDA lot 1 (data not shown).

#### Precision

Assay imprecision was estimated from two data sets. First, two human serum specimens from MPXV-infected subjects and three human serum specimens from Jynneos-vaccinated subjects were selected. Each specimen was tested in duplicate, with each replicate tested utilizing two wells, across six separate days (Figure S5A). These specimens spanned a broad range of antibody concentrations within the assay’s quantifiable limits, as determined by calibrator performance. Second, results from testing the manufacturer supplied serology controls (1,2, and 3), with two technical replicates (two wells), over ten days were also analyzed (Figure S5B). Collectively, the three controls span the assay’s quantitative range. Control 3 fell just below the lower limit of quantification (LLoQ) for MPXV A29L, VACV A27L, and MPXV B6R antigens, which is expected since control 3 is presumed negative. Variance decomposition was performed using ANOVA to estimate intra-assay, inter-assay, and within-laboratory components. Results with an inter-well geometric coèicient of variation (GCV) > 37% were excluded, as this level of variability between technical replicates may indicate an unreliable measurement. Only human serum specimen V4 was àected, with one VACV L1R result (day 1) and one MPXV M1R result (day 10) removed. All human serum specimens demonstrated within-laboratory imprecision with GCV <37% (Figure S6A). For the serology controls, despite some control 3 results falling below the LLoQ, all three controls exhibited within-laboratory imprecision with GCV < 37% (Figure S6B). Notably, the serology controls showed lower and more consistent imprecision compared to the selected specimens described above, so they don’t appear to entirely reflect real-world variability.

#### Accuracy

Accuracy was evaluated by comparing measured results from the manufacturer-supplied serology controls (Control 1, Control 2, and Control 3) to their manufacturer assigned AU/mL values. Control 1 and Control 2 are expected to fall within the high and mid quantitative ranges of each assay (Figure S5B). Each control was tested in duplicate wells across ten independent runs. For both Control 1 and Control 2, the relative error compared to the manufacturer-assigned AU/mL values was within ±30% for all assays (Figure S7A). Control 3 serves as a presumptive negative control, with assigned AU/mL values of < 0.1 AU/mL for VACV A27L, MPXV A29L, VACV A33R, MPXV A35R, VACV B5R, MPXV B6R, VACV D8L, and MPXV E8L, and < 0.2 AU/mL for VACV L1R and MPXV M1R. Control 3 was tested alongside the other controls in duplicate wells across ten runs (Figure S5B). The geometric mean response and geometric standard deviation (GSD) were calculated, and an empirical limit for Control 3 was determined by adding three times the GSD to the geometric mean (limits shown in red in Figure S7B). All empirically determined limits for Control 3 were ≤ 0.1 AU/mL. Together, these results demonstrate the accuracy of the assay relative to the manufacturer-supplied serology controls (Controls 1–3) since the percent relative error of control 1 and 2 are < ± 30% and Control 3 AU/mL values were ≤ 0.1 AU/mL.

#### Analytical Measurement Range

The lower limit of quantification (LLoQ) was defined as the minimum concentration above the limit of detection (LoD) that meets precision criteria and falls within the assay’s linear range. The upper limit of quantification (ULoQ) was defined as the maximum concentration that meets precision criteria and remains within the linear range. Together, the LLoQ and ULoQ establish the analytical measurement range (AMR) of the assay. All calibrator-derived results within the LLoQ and ULoQ met acceptance criteria for both linearity (Figure S4) and imprecision (Figures S5 and S6). Therefore, the LLoQ and ULoQ values presented in Table S1 define the assay’s validated AMR.

#### Robustness

Since clinical specimens are likely to undergo several freeze thaws (FT) between collection and testing, the èect of FT on the assay results was evaluated. Four specimens were selected, two MPXV-infected (M42, M48) and two smallpox vaccinated (V14, V57). Each specimen was thawed and four aliquots prepared and frozen back down. One tube was designated as freeze-thaw (FT) 0 and not thawed again till testing, one tube was designated as FT 1 and was thawed once and frozen down again prior to testing, one tube was designated as FT 2 and was thawed and frozen twice prior to testing, one tube was designated as FT 3 and was thawed and frozen three times prior to testing. The ratio of antibody level results between FT 0 and each of the dìerent freeze-thaw specimens (1–3), was calculated and all ratios were within 0.5 -1.5-fold, so no significant freeze-thaw èects were detected (Figure S8).

---

**Figure S1:**
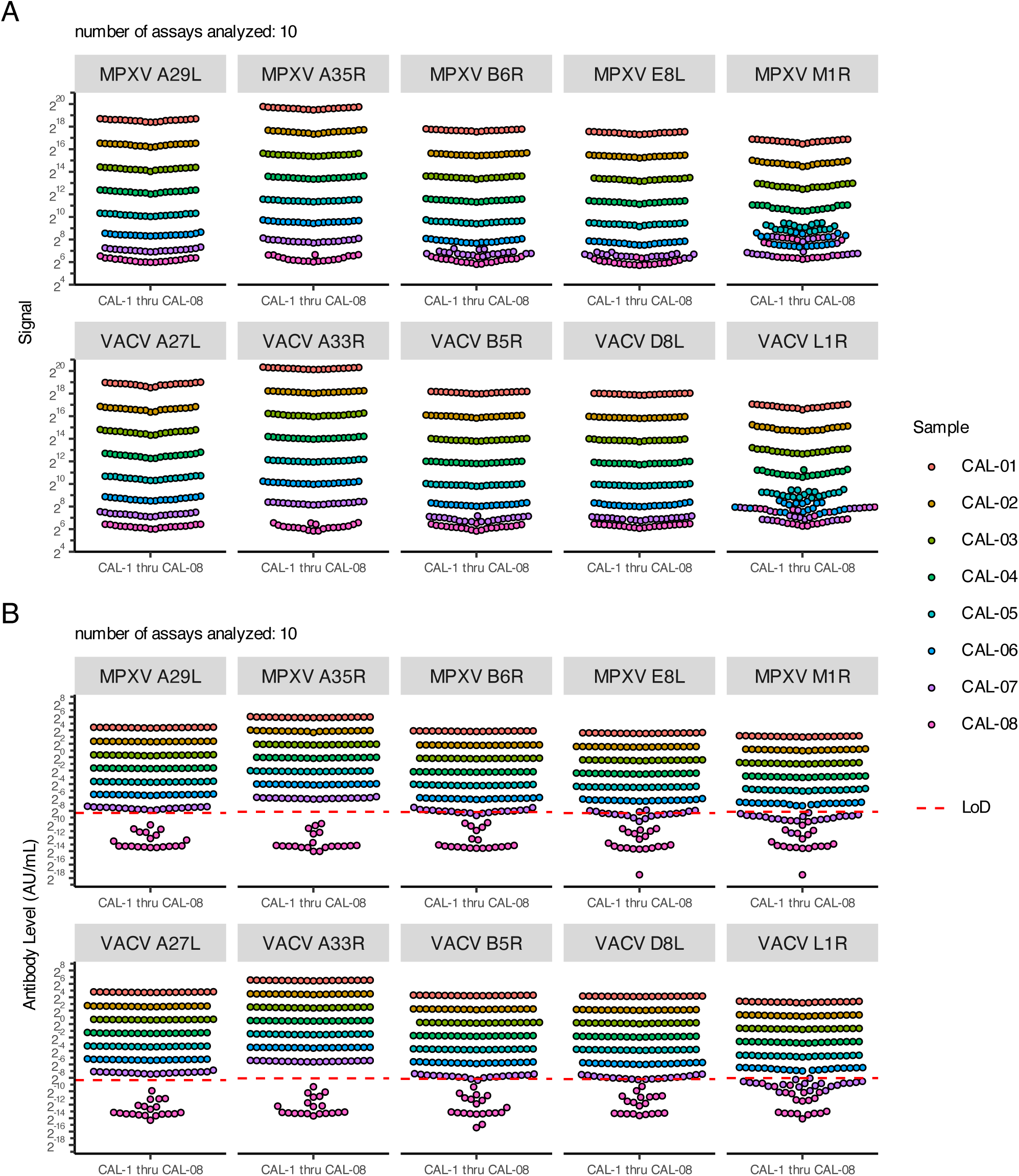
Signal results from different days overlap among the lowest calibrators, but conversion to AU/mL normalizes for the day-to-day differences in Signal. Shown are calibrator results from ten independent runs of the MSD Orthopoxvirus assay in units of Signal (A) and antibody level in AU/mL (B). For MPXV A29L, VACV A27L, MPXV A35R, VACV A33R antigens CAL-01 thru CAL-08 are entirely distinguishable by Signal even between runs on different days. However, the remaining antigens showed some degree of Signal overlap between CAL-05 thru CAL-08. Examination of Signal levels within any given run showed that the Signal levels are distinguishable for CAL-01 thru CAL-07 for each of the antigens. Conversion of Signal to AU/mL normalizes results such that overlap is limited to CAL-07 and CAL-08. The limit of detection (LoD) for each antigen for each run was calculated following the described manufactures recommendation and the average of these results is indicated by the red dashed line. When the CAL-08 signal fell below the lower asymptote of the 4PL model used to convert Signal to AU/mL, an AU/mL value could not be calculated and was arbitrarily set to the lowest reportable value based on the 4PL model fit.

**Figure S2:**
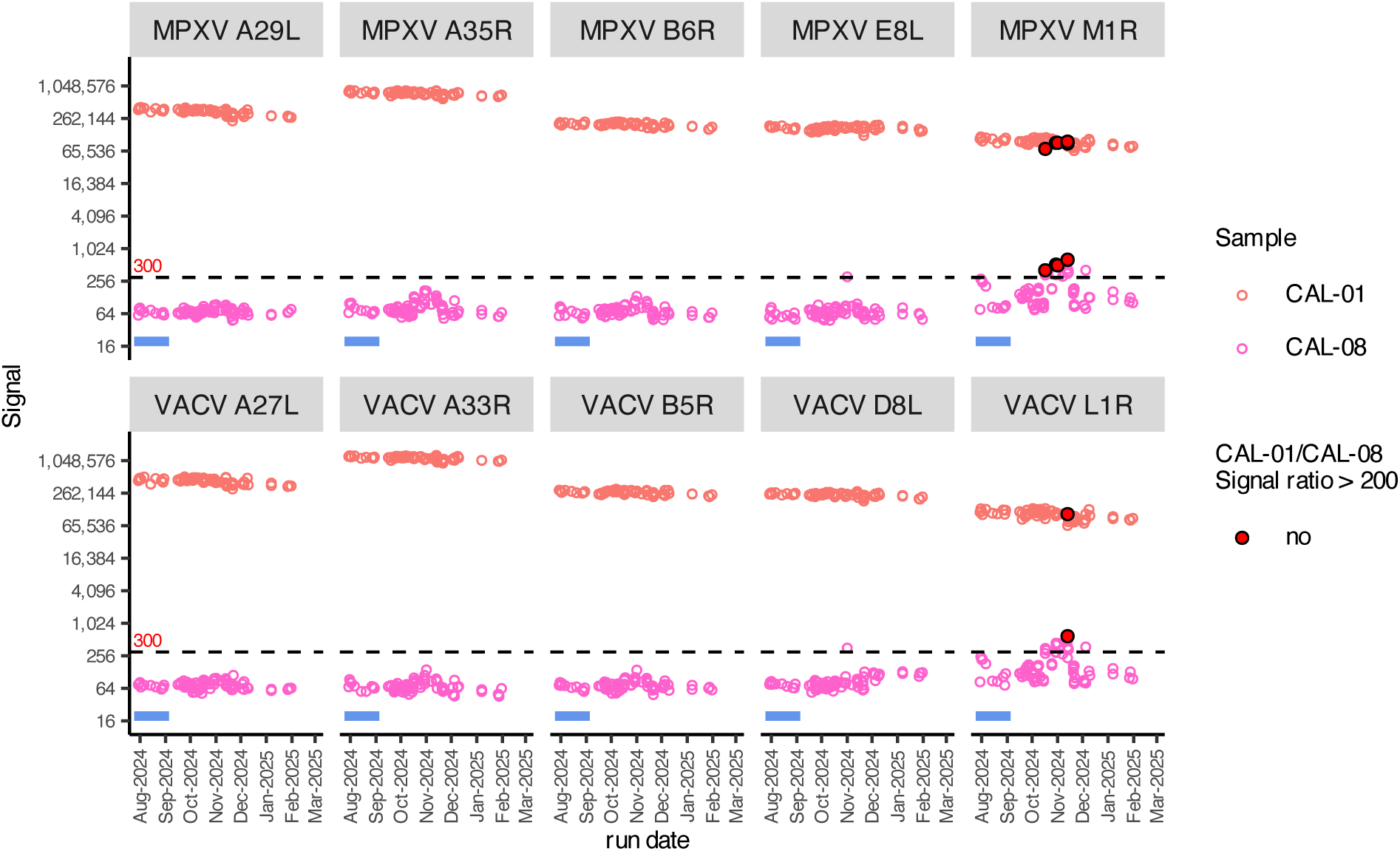
Greater variation in CAL-08 Signal over time is evident for antigens MPXV M1R and VACV L1R. Shown are CAL-01(red) and CAL-08 (purple) Signal values both during validation of the assay (see light-blue shaded bars) and subsequent testing over the following 5-6 months. The dashed blank line indicates the initial quality control threshold that CAL-08 was expected to not exceed. Red filled points are assays that did not meet CAL-01/CAL-08 Signal ratio of at least 200, which included five M1R assays and one L1R assay.

**Figure S3:**
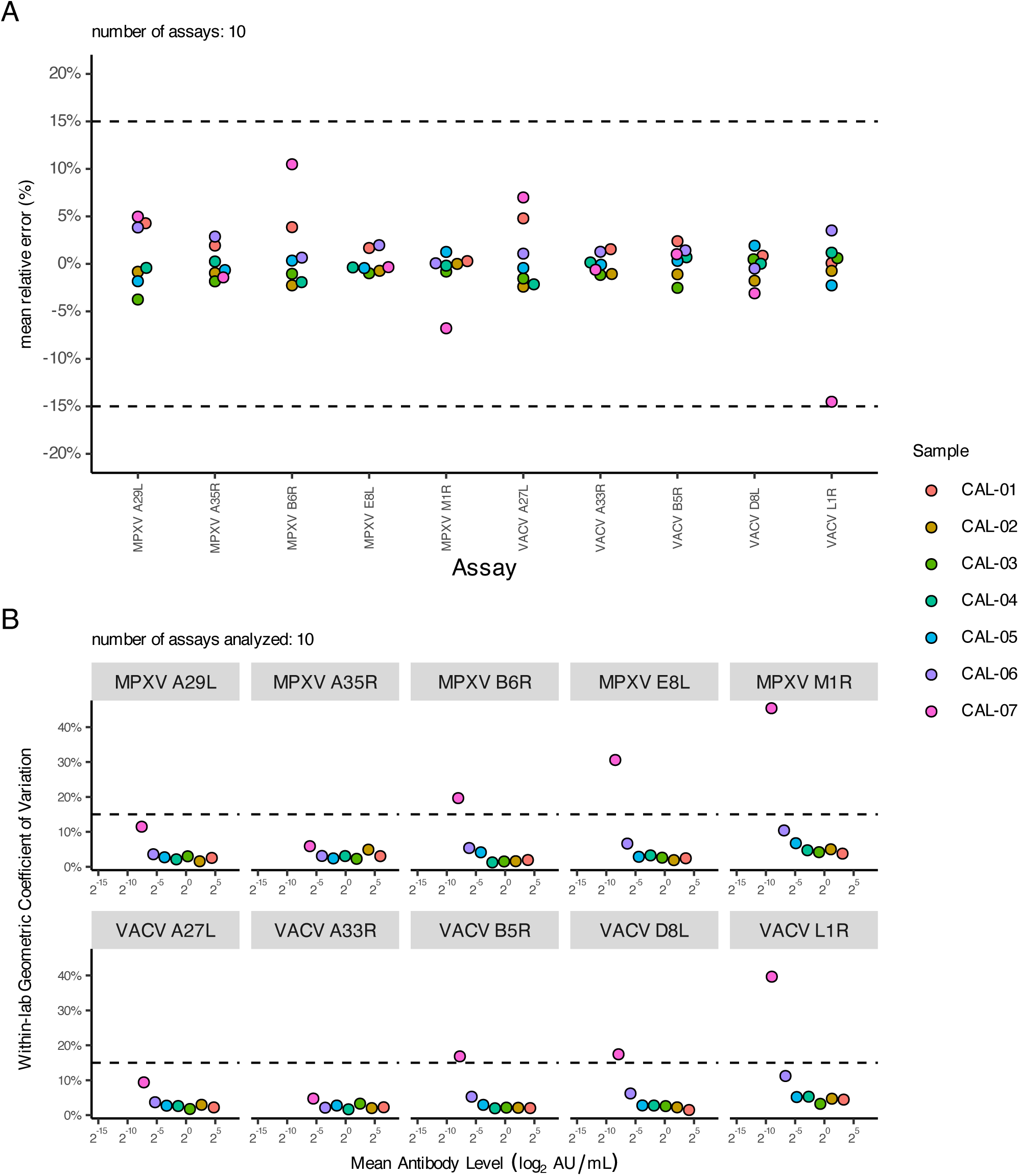
Average relative error of calibrators is within +/- 15 and within-lab geometric coefficient of variation of each calibrator is within 15% for CAL-01 thru CAL-06. Assay calibrators (CAL-01 thru CAL-07) were tested in duplicate over ten runs (A) Shown is the mean percent relative error for each calibrator (CAL-01 thru CAL-07) with the acceptability criteria of mean relative error of +/- 15% indicated by black dashed line. The mean relative error estimates are based on AU/mL values. (B) From the same data the within-lab geometric coefficient of variation (GCV) was calculated and plotted for each calibrator. Dashed black lines mark the acceptability criteria of a within-lab GCV of 15% or lower. Both the mean relative error estimates and the %GCV is calculated from AU/mL values.

**Figure S4:**
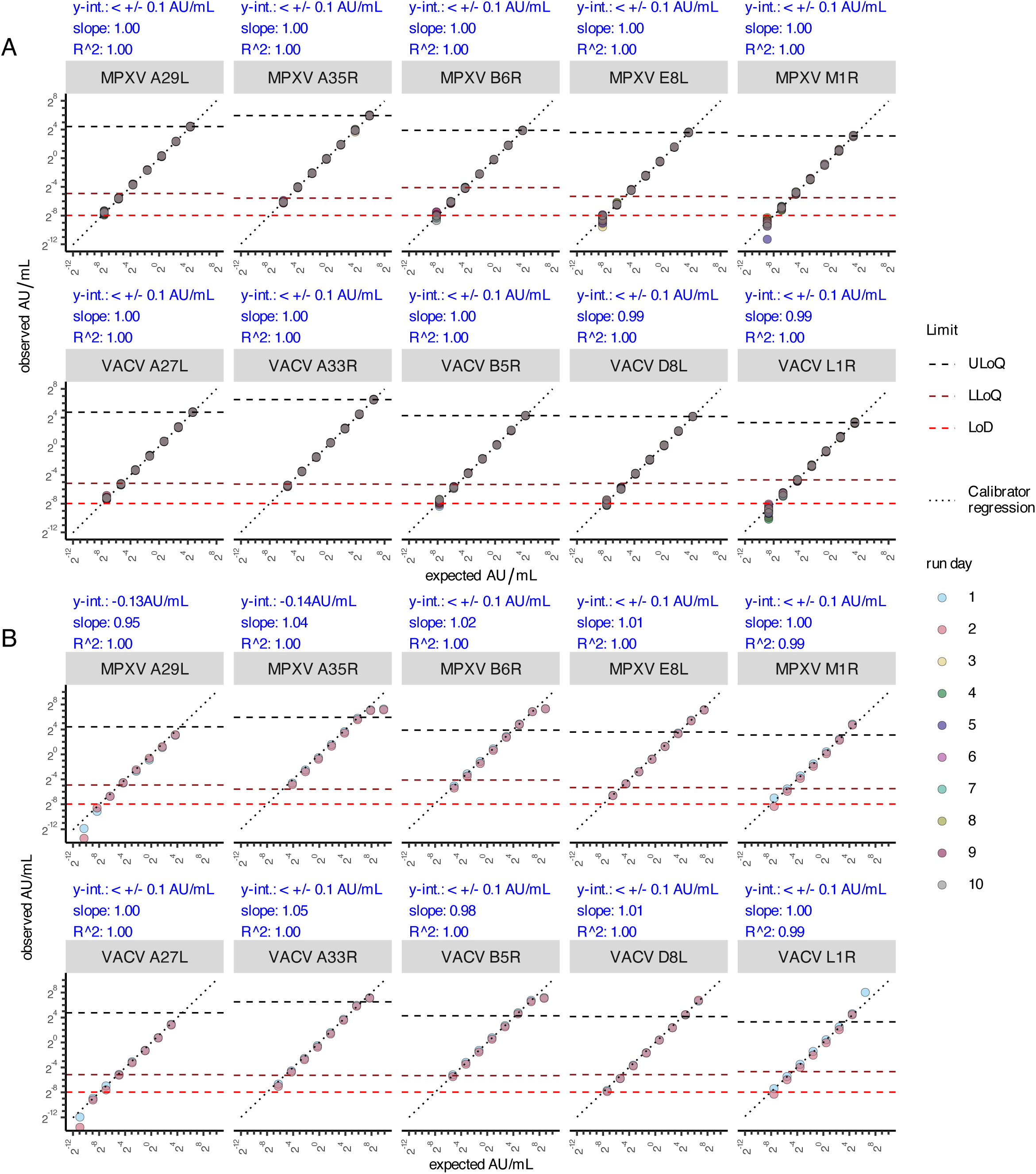
Antibody levels in the quantitative range of the MSD Orthopoxvirus assay are strongly linear. Linear regression was performed against observed (y-axis) and expected (x-axis) antibody levels (AU/mL) from testing (A) assay calibrators over ten days or (B) serially diluted serum specimen from MPXV-infected individual over two days (M32). Only results within the lower and upper limits of quantification were considered for linear regression (see Table S1). The expected AU/mL values for the calibrator are based on values provided by the manufacturer and the expected AU/mL values for the M32 specimen were based on testing of the M32 specimen in the MSD Orthopoxvirus assay at a dilution of 1/5000. The y-intercept, slope and R2 value determined from each linear regression is summarized in blue text above each plot. The dotted line represents the best fit line based on the linear regression of the calibrators only. The LoD, LLoQ, and ULoQ for each antigen are indicated in red, dark red, and black dashed lines, respectively.

**Figure S5:**
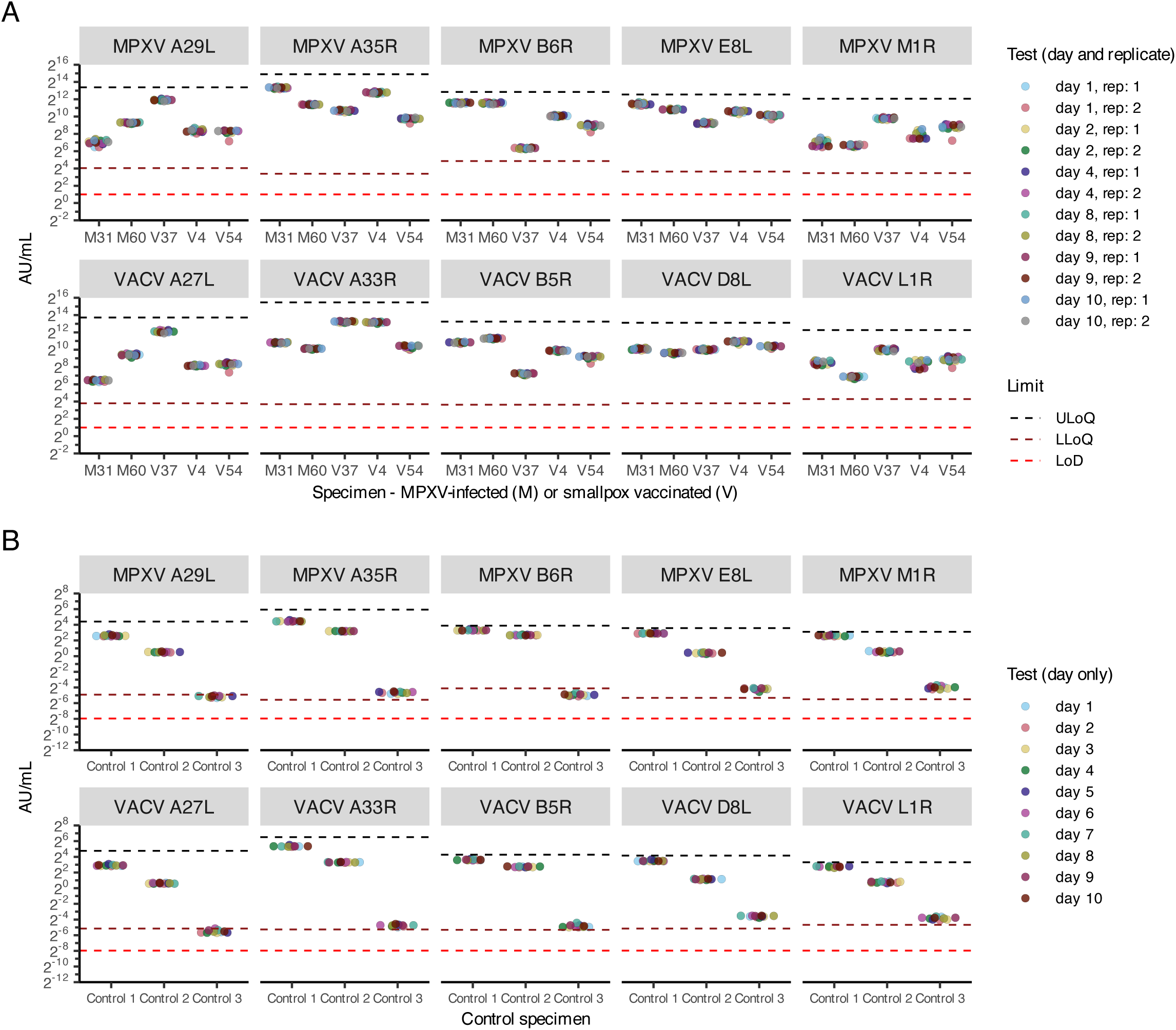
Antibody level (AU/mL) results for estimation of assay imprecision from either human serum specimens or serology controls provided by the manufacturer span the quantitative range of the assay. (A) Two serum specimens from MPXV-infected subjects (M) and three serum specimens from vaccinated subjects (V), which represent a range of antibody levels in the assay were selected for assessing assay imprecision. Specimens were tested over six days in duplicate, with reach replicate tested with two technical replicates (i.e. two wells per replicate) for a total of twelve average results per specimen. (B) Assay controls were tested once per run and the ten results shown are the average response from two wells. The ULoQ (black dashed line), LLoQ (brown dashed line), and LoD (red dashed line) established based on calibrator performance are shown. The LoD, LLoQ, and ULoQ are higher in (A) than (B) since human serum specimens were tested at a 500-fold dilution, whereas the serology controls were tested neat.

**Figure S6:**
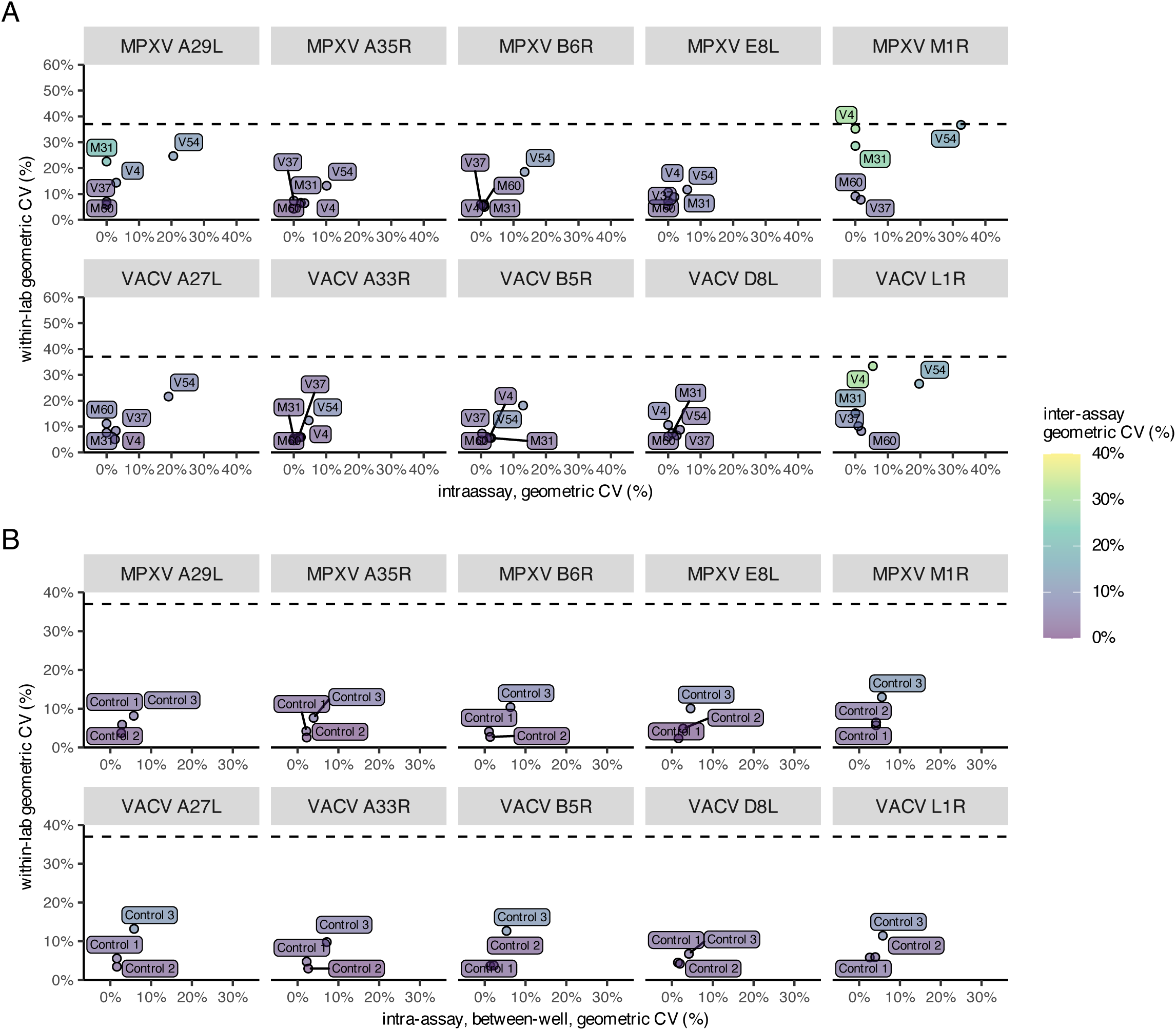
Imprecision of human serum specimens and serology controls meet acceptance criteria of within-lab geometric coefficient of variation (CV) of < 37%. Based on the testing shown Figure S5, variance decomposition using ANOVA was performed to determine within-lab (y-axis), inter-assay (coloring of labels and points), and intra-assay imprecision (x-axis). (A) Are the imprecision results from testing human serum specimens shown in Figure S5A. Specimens IDs include whether there are from individuals MPXV-infected (M) or smallpox vaccinated (V). For specimen V4, a day 1 VACV L1R result and a day 10 MPXV M1R result, both with between-well geometric CV of greater than 37%, were removed for this analysis. (B) Are the imprecision results from testing the serology controls. Since the serology controls were only tested once, with two technical replicates, the intraassay imprecision is between-well.

**Figure S7:**
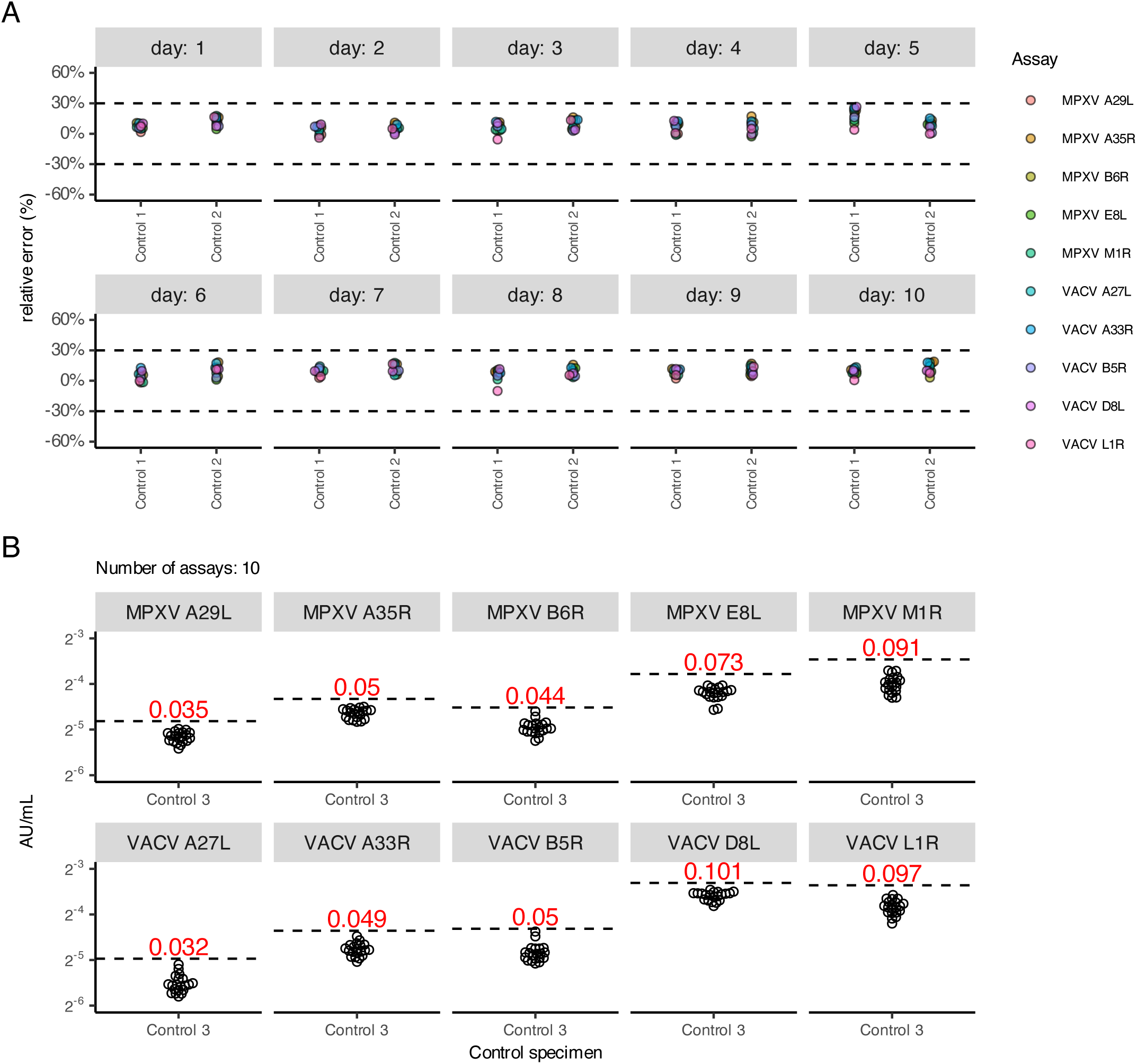
Based on manufacturer supplied serology controls the MSD Orthopoxvirus assay is accurate given that day-to-day relative error for Control 1 and 2 is less than +/- 30% and control 3 limit antibody levels are all less than 0.2 AU/mL. Serology controls (1,2 and 3) provided by the manufacturer were tested with two technical replicates over ten days. (A) Shown are the results for serology controls 1 and 2. The dashed black lines mark the acceptance criteria of a relative error of +/- 30%. (B) Shown are the results for serology control 3. The dashed black line is set to the control 3 geometric mean plus three times the control 3 geometric standard deviation. The acceptance criterion is meet since the Control 3 limit is less than 0.2 AU/mL for each antigen.

**Figure S8:**
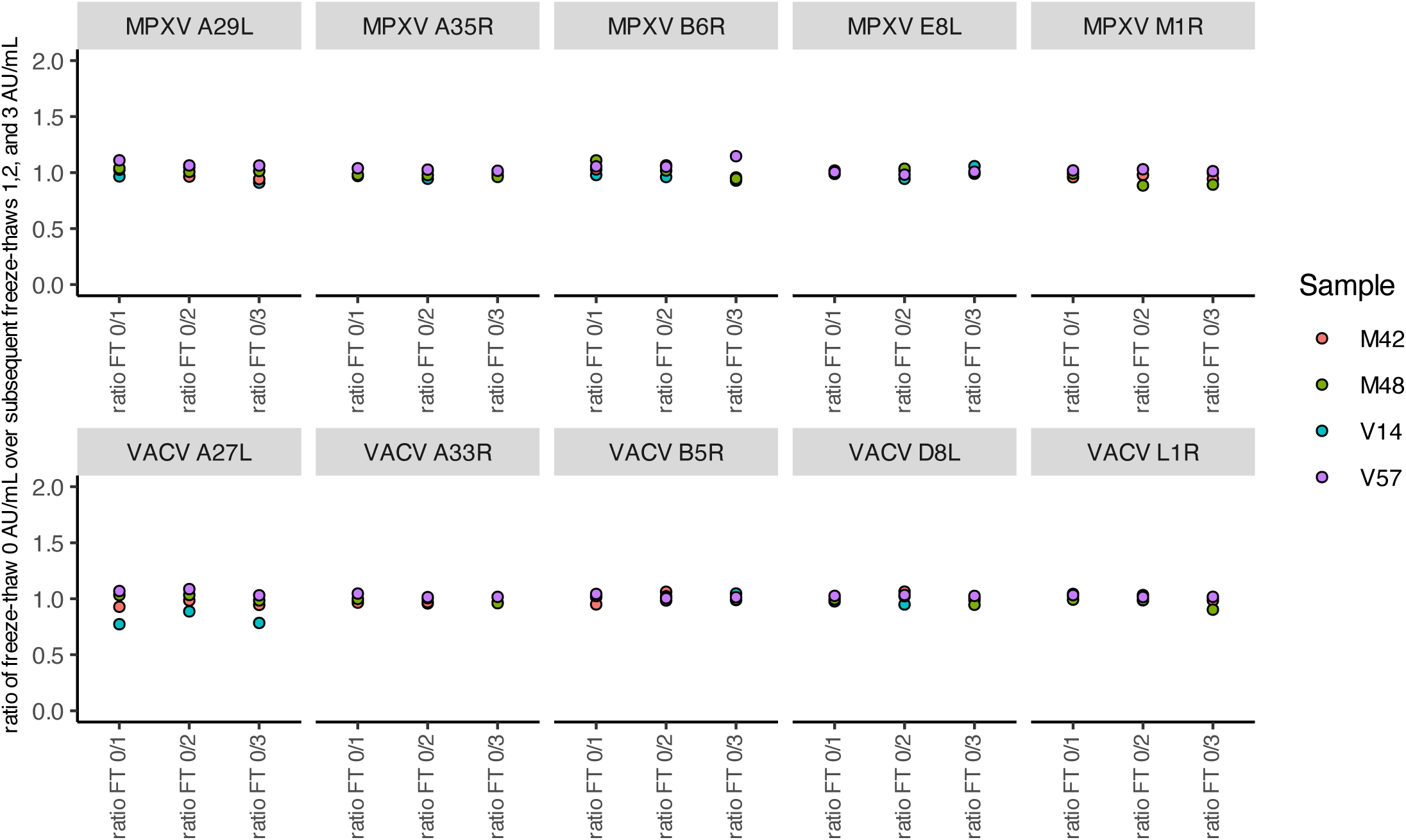
The assay is robust against freeze-thaw cycles of serum specimens. Plotted is the AU/mL ratio of the results before freeze-thaw (FT 0) over results from freeze-thaw 1 (ratio FT 0/1), from freeze-thaw 2 (ratio FT 0/2), or from freeze-thaw 3 (ratio FT 0/2). Shown are results from testing four specimens and specimens IDs include whether there are from individuals MPXV-infected (M) or smallpox vaccinated (V).

**Figure S9:**
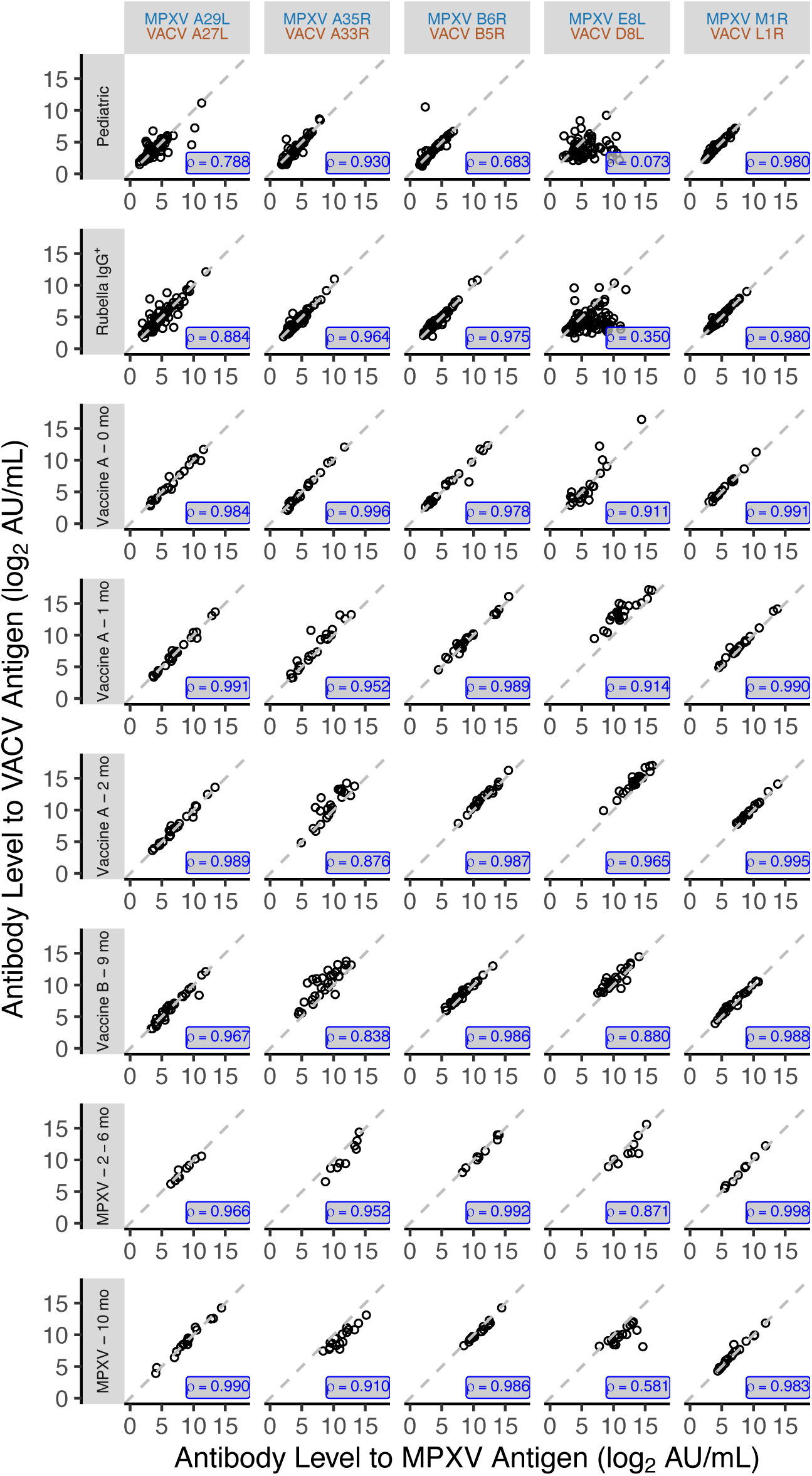
Antibody titer results against antigen orthologs are highly correlated. Shown are pairwise plots of the log2 transformed AU/mL results between antigen ortholog pairs within in each cohort. Inset value in blue is the Pearson’s correlation coefficient, also shown in Figure 1A. The P61 MPXV B6R/VACV B5R results were omitted from the plot, since the VACV B5R antibody level was undetected.

**Figure S10.**
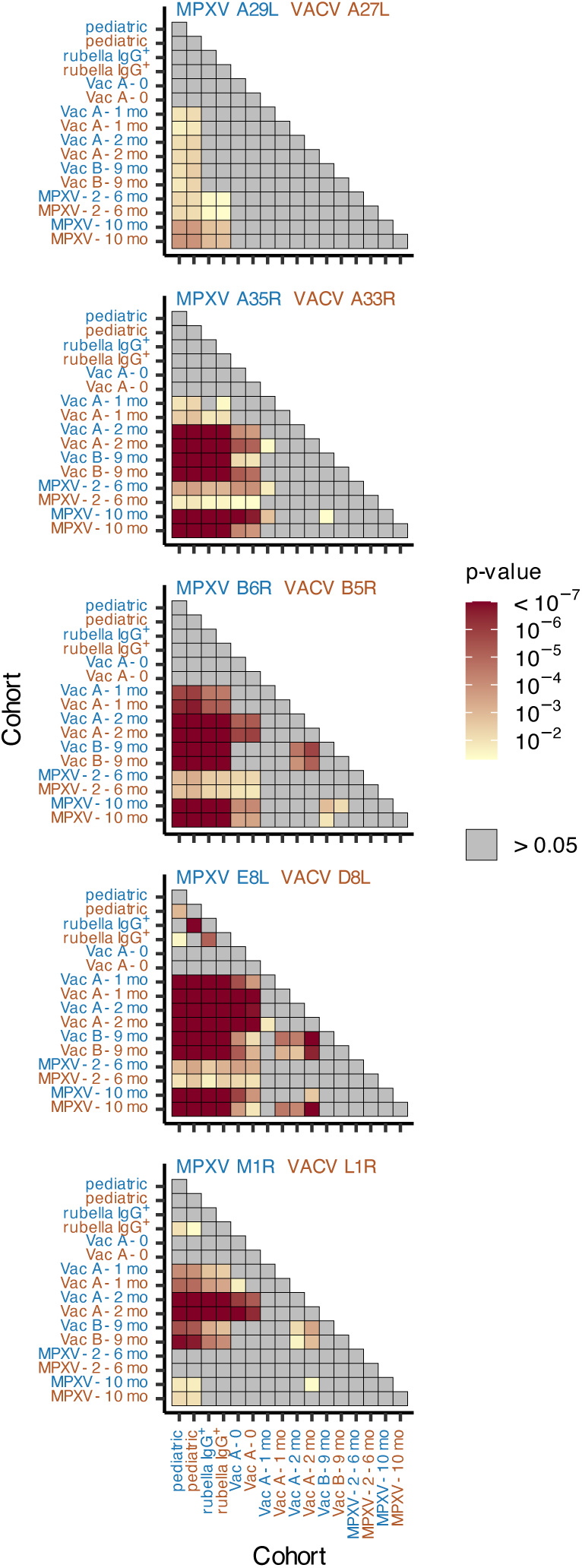
Significant differences in anti-orthopoxvirus antibody levels among MPXV-infected, vaccinated, and negative control cohorts. For each MPXV and VACV antigen pair (indicated along the top of each matrix), log-transformed antibody levels (AU/mL) were compared between cohorts using two-sided t-tests. Cohort names are color-coded: blue for MPXV antigen results and brown for VACV antigen results. The p-values are displayed in a lower-triangular matrix, where each tile represents a pairwise comparison as indicated by the intersecting column and row names. Tile shading intensity is inversely proportional to the p-value (i.e. darker red corresponds to a smaller p-value); gray tiles indicate p-value > 0.05. All p-values are Bonferroni-adjusted for multiple comparisons.

**Figure S11:**
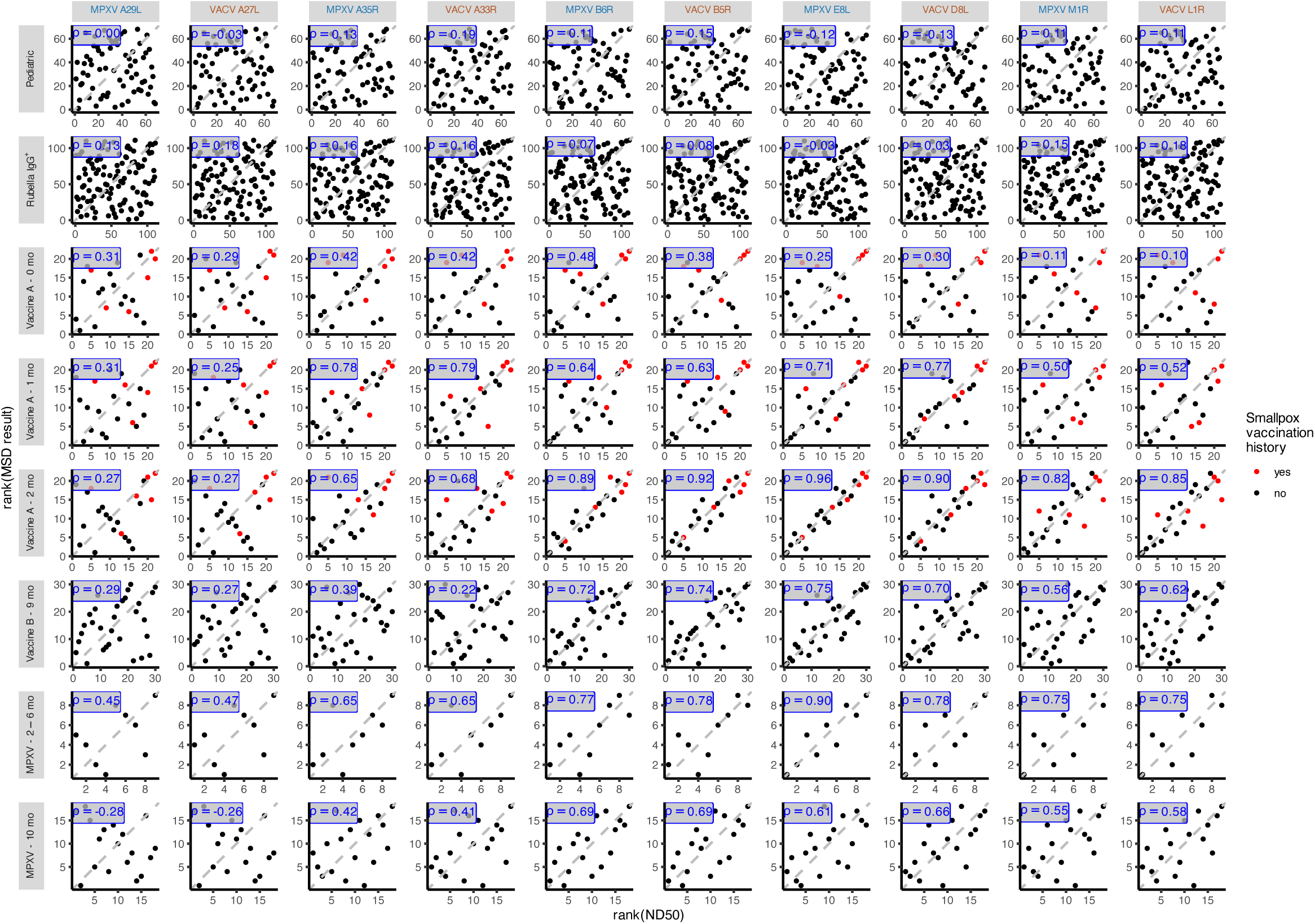
The anti-orthopoxvirus antibody titer results and MVA neutralization results are generally correlated in sera from individuals know to be MPXV infected or vaccinated. Shown are pairwise plots of the rank ordered MSD AU/mL results (MSD Response) verses rank ordered MVA neutralization assay ND50 results. Inset value in blue is the Spearman’s correlation coefficient, also shown in Figure 4B. Results are filled with red if specimen is from an individual with known smallpox vaccination history.

**Figure S12:**
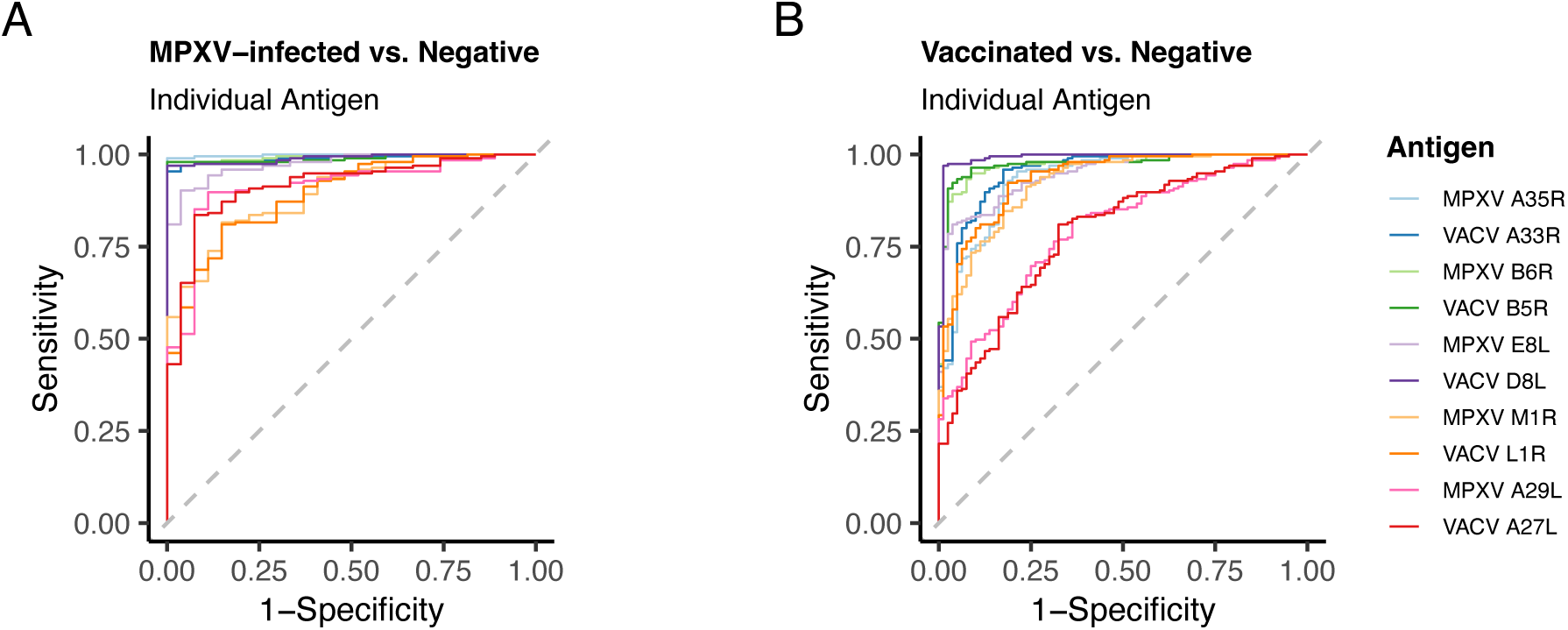
Receiver operator characteristic (ROC) curves separately comparing each exposed group (MPXV-infected or vaccinated) to negative controls. MPXV-infected refer to all sera from known MPXV infected individuals, Vaccinated refers to all sera from known vaccinated individuals, and Negative refer to all sera from the rubella and pediatric negative cohorts combined. Baseline sera collected prior to vaccination (Vaccine A – 0 month) were included in the Vaccinated group if they had a history of smallpox vaccination, otherwise they were categorized as negative (6 out of 22 individuals had previous history of vaccination). (A) ROC curves for all individual antigens generated by comparing all known MPXV-infected (n = 27) to all negatives (n = 195). (B) ROC curves for all individual antigens generated by comparing all known Vaccinated (n = 80) to all to all negatives (n = 195). Summary statistics from this analysis can be found Tables S5-S6.

**Figure S13:**
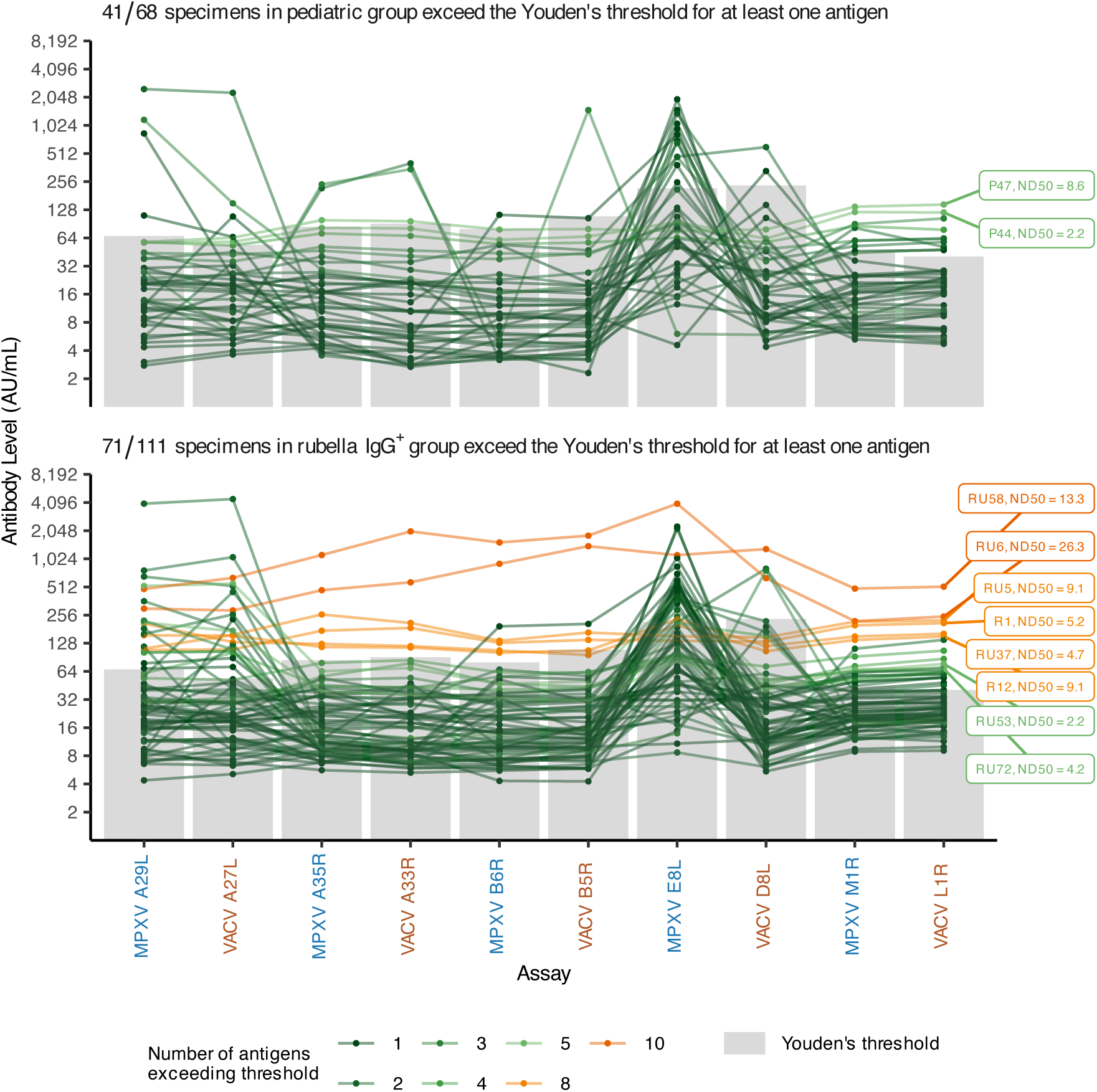
Some specimens in the negative cohorts (pediatric and rubella IgG+) exceed Youden’s threshold (based on distinguishing exposed and unexposed groups) for one or more antigens. For each negative cohort (upper, pediatric; lower, rubella IgG+) the antibody level (AU/mL) is plotted for specimens that exceed Youden’s threshold based on distinguish exposed vs. unexposed groups for at least one antigen (see Table S5). Specimens that exceeded the threshold for five or more antigens are labeled with the specimen identifier and MVA-neutralization ND50.

